# Mathematical modeling suggests pre-existing immunity to SARS-CoV-2

**DOI:** 10.1101/2021.04.21.21255782

**Authors:** Marcus Carlsson, Gad Hatem, Cecilia Söderberg-Nauclér

## Abstract

Mathematical models have largely failed to predict the unfolding of the COVID-19 pandemic. We revisit several variants of the SEIR-model and investigate various adjustments to the model in order to achieve output consistent with measured data in Manaus, India and Stockholm. In particular, Stockholm is interesting due to the almost constant NPI’s, which substantially simplifies the mathematical modeling. Analyzing mobility data for Stockholm, we argue that neither behavioral changes, age and activity stratification nor NPI’s alone are sufficient to explain the observed pandemic progression. We find that the most plausible hypothesis is that a large portion of the population, between 40 to 60 percent, have some protection against infection with the original variant of SARS-CoV-2.

## 1 Why do SEIR-models fail, or do they?

For most countries or regions, when observing the time series of new cases, hospitalizations or deaths by COVID-19, it is clear that they go up and down in a manner that can only be explained by Non-Pharmaceutical Interventions (such as lock-downs, school closures etc.), seasonal and behavioral changes. However, in a few places where the spread got out of control, such as Manaus and New York, the curves look very much like the “wave of infections” predicted by standard mathematical models for infectious diseases, called SEIR. In particular this applies to Stockholm where the authorities have been unwilling to enforce any major NPI’s, and the recommendations (as well as other key parameters such as season) have been virtually constant, especially during the second wave.

Despite this, the measured sero-prevalence in all to us known locations (also towns such as Bergamo [52]) is well below the mathematically predicted herd-immunity threshold, which is estimated to somewhere above 60%. The city of Manaus had a sero-prevalence below 30% by October [13], yet despite the society being relatively open it was not hit by a second wave until late December, most likely caused by a mutant virus variation [4]. The sero-prevalence in New York after the first wave was 23% [6] and a measurement from Stockholm indicates that 22% of the population had had the virus in February, right after the second wave.

Is it possible that this discrepancy between the SEIR model and reality is only due to variations in the effective *R*-value caused by changing NPI’s, or is the SEIR-model simply inapt for modeling COVID-19? Or could we refine the SEIR-model to give more realistic output? Or could it be that the parameters we feed the model are wrong? This is the question we tackle in this paper, whose supplementary material contains a thorough revision of heterogeneous SEIR models and their parameters.

We argue that the first three explanations are less credible, and then revisit the idea of a pre-existing immunity to SARS-CoV-2, or pre-immunity for short. We do not address the nature of this protection, which either can stem from pre-existing cross-reactive adaptive immunity or different degrees of innate immunity, but we mention the publications [18, 43] showing different mechanisms where prior exposure to other viruses can prevent infection by SARS-CoV-2. We also point to our own paper [15], showing that pre-existing immunity could in fact be a mathematical manifestation of great variability in susceptibility to SARS-CoV-2. Hence the results of this paper do not necessarily imply that some people have sterilizing immunity, but rather that a large proportion have a relatively low risk of getting COVID-19 when exposed to moderate dose of SARS-CoV-2. To avoid confusion with sterilizing individual pre-immunity, we call the latter type of immunity “population pre-immunity”.

We show that upon taking population pre-immunity into account, it is possible to accurately model both the first and second wave of COVID-19 in Manaus and Stockholm. It is our hope that this work can help improve the understanding of this new virus and stimulate medical research aiming to further explore potential causes for pre-immunity. This paper was written before the third wave hit, and another paper which extends the model to include the third wave is found here [14].

## 2 A close look at the SEIR-setup

The mathematical models recommended for respiratory diseases like influenza are (refinements of) a basic ordinary differential equation commonly known as SEIR, (see e.g. Ch. 4 and 9, [10]). While these deterministic models do not capture the random nature of real life, it has been shown that more advanced stochastic models behave like these deterministic models when there is a major outbreak in a large population (see e.g. [3] Ch. 5.5 and [7]).

In the acronym SEIR, *S* = *S*(*t*) stands for the amount of susceptible at time *t* in a population of *N* individuals, and *s*(*t*) is the corresponding fraction of the population in group *S*, so *s*(*t*) = *S*(*t*)*/N* equals 1 at the beginning of the pandemic (unless there is pre-existing immunity, easily included by simply choosing an initial value less than one). *E*(*t*) stands for exposed, to account for the incubation time. *I*(*t*) stands for infectious, after which people recover and “appear” in *R*(*t*) instead. Like *s*, the letters *e, i, r* will denote the respective fractions of the population, so if *i*(*t*) = 0.02 it means that 2% of the population is infective on the particular day *t*. The majority of individuals will thus start in *s* and end in *r*, but not all. The final value of *r* is called the final size of the epidemic, usually a bit higher than the herd-immunity threshold because the *r*-curve overshoots due to the momentum it gets from having a large fraction in *i* at the same time. The equation system depends on a few additional parameters, most notably the *R*_0_ value that determines the speed of transmission, but also the mean incubation time *T*_*incubation*_ and the mean infectious period *T*_*infectious*_, explained in depth in the Supplementary Material (SM), Section 5 and 5.1.

An important issue to stress is the difference between the curve *i*, which models all infectives at a given moment *t*, and the fraction of *new infections* on a given day, which equals *ν*(*t*) := *s*(*t* − 1) − *s*(*t*). These two are often confused and the latter curve is usually much smaller than the former (since people remain infective for some days but only become infected once). Measured curves for new infections based on PCR or antigen-testing will scale linearly with *ν*, not with *i*.^1^

### 2.1 Discrepancies between model and reality

Simple models such as SEIR rely on gross simplification of human life, but these may still be useful if their *main characteristics* correlate acceptably well with reality. By main characteristics we mean quantities such as the time *T*_*wave*_ it takes for the outbreak to pass, around 70 days in Figure 1, which is roughly the time observed in Manaus (for both waves). Another key quantity is the *final size of the epidemic* i.e. the fraction of people who at some point had COVID-19, which we define as 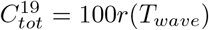. In Figure 1, 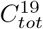 is close to 80% and in the right graph we see that a whopping 2.5% will fall ill on the same day! To our knowledge this behavior is not near reality in any hard hit location of the planet (including naval ships and prisons), and yet, this is what many qualified experts and proponents of the mitigation-strategy had predicted would happen in early 2020 (see e.g. [26]). So why does the model fail so drastically?

**Figure 1:**
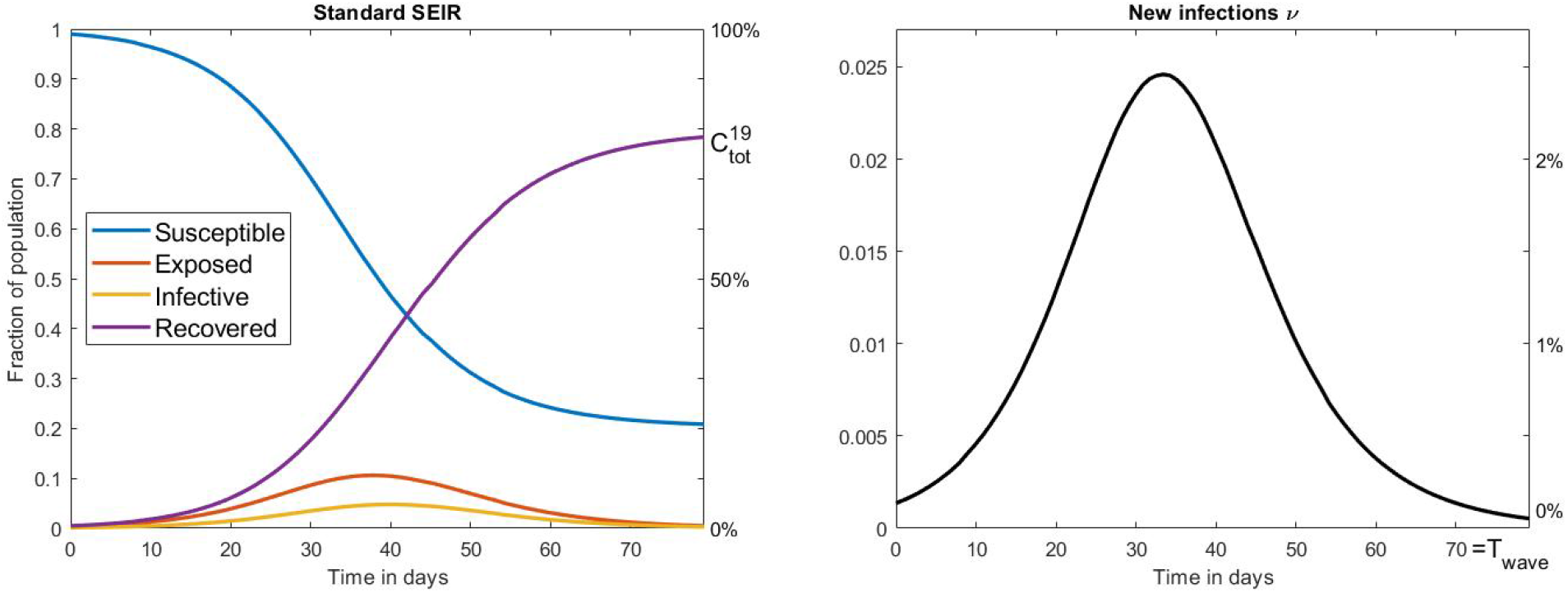
Left: SEIR outcome with *R*_0_ = 2. Right: New infections *ν*.

One answer could of course lay in the (overly simplistic) setup of the equation system, as argued e.g. in [16]. We review the setup of basic SEIR in the Supplementary Material (Sections 5-5.1) with the aim of tuning the model to COVID-19. In short summary; on one hand there is a big gap between the SEIR-model and reality, and parameter choices are somewhat ad hoc. On the other hand, the model is fairly robust and different parameter choices do not affect the overall outcome drastically. Moreover, more accurate equation systems that also take the age of infection into account give almost identical output as the SEIR-equation system (see in particular the blue vs. the yellow curve in SM Figure 8), and hence it seems that these shortcomings can not explain the discrepancy between SEIR-based predictions and observations.

It is important to realize that in theory, these models are valid also under NPI’s, as long as these are held constant. For then there will be an artificial fixed *R*_0_ value that is adjusted for the corresponding NPI’s, which we will denote by 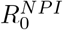. So lets try to lower 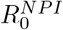 so that 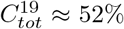 to match the observed data from Manaus [13]; 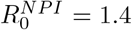 does the job, see Figure 2. However, now the wave is expected to be around 150 days, twice the time suggested by real data. Moreover, there are indications that the measured sero-prevalence at peak in Manaus most likely did not reflect the sero-prevalence in society, which probably was closer to 30% [30]. If this is the case, then the SEIR-model is off even more drastically.

**Figure 2:**
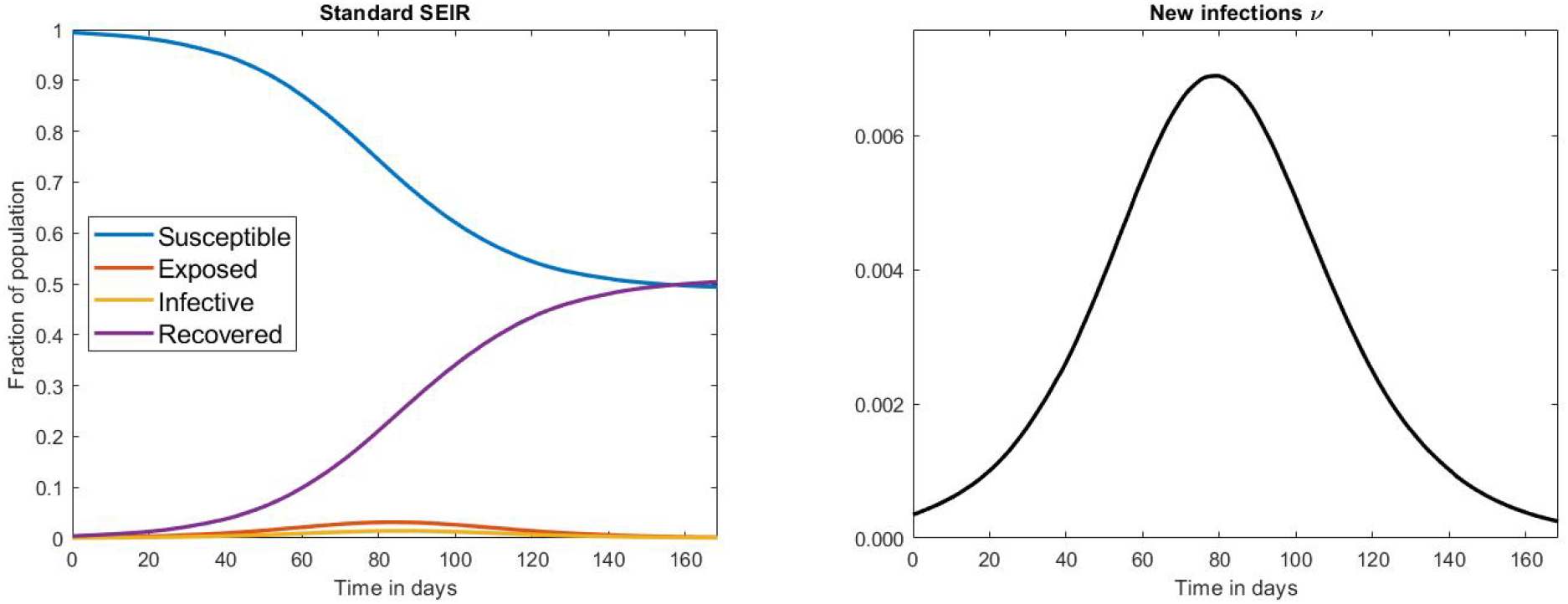
Left: SEIR outcome with *R*_0_ = 1.4. Right: New infections.

The Manaus curve is strikingly similar (in shape and duration) to the one for New York, which landed at 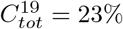 [6] as well as the one for Stockholm (that did not go into lock down) and measured a sero-prevalence of 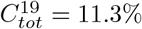 in June [23]. With SEIR-modeling these outbreaks should have taken 1 and 2.5 years respectively, and is hence not realistic. This behavior is easy to understand on an intuitive level; in order for the final size of the epidemic to be small the 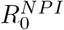 must be near one and then the virus spread becomes unrealistically slow. Thus, the problem with SEIR-models is that at least one of the key output-parameters *T*_*wave*_ and 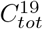 will be totally at odds with reality. *In other words, no matter how you tune the input parameters, the model generates curves which either miscalculates the pace or the magnitude of the epidemic outbreak*.

### 2.2 Possible explanations

A quick fix to the above dilemma is to include pre-immunity. In the SM Section 5.2 we show that for every solution to the SEIR equation system in the absence of pre-immunity, one can obtain a solution with a level *θ* of pre-immunity simply by multiplying the solution by the constant factor (1 − *θ*) and adjusting *R*_0_ accordingly. In other words, if we have found a solution which fits a given time-series well except for the magnitude, we may simply rescale it. To be concrete, if a model has a suitable shape and predicts *T*_*wave*_ correctly, but gives 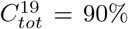 whereas measured sero-prevalence indicates 52%, this could indicate that 42% are actually immune to infection, since 52 ≈ (1 − 0.42) · 90. We investigate this possibility in Section 3, but first we will discuss other potential explanations.

It can be argued that the type of stochastic models mentioned above are not apt for modeling of SARS-CoV-2, which indeed is a peculiar virus that spreads in clusters, and it is estimated that 80% of the cases are caused by less than 20% of infected individuals, the so called “super-spreaders” [1, 21]. In Section 7.3 we show that, rather surprisingly, adding this complexity to the deterministic SEIR-model does not in any way alter the output. It could then be argued that one needs to take randomness into account, which has been done in [25]. They consider a stochastic SEIR-model where *R*_0_ is a random variable (for each infected individual) with a “fat-tailed” distribution, and while this displays an erratic and possibly more realistic behavior, its effect on *T*_*wave*_ and 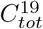 still does not seem to overcome the shortcomings discussed above (see in particular Figure 4 in [25]).

It has been argued that population heterogeneity may explain the gap between model and observation, see e.g. [12] and [53]. In Section 6 we take a closer look at these and show that while heterogeneity certainly plays an important role for more accurate modeling, its effect on model outcome is not substantial enough to explain the huge gap between model and observation. More precisely, when applied to the data series from Stockholm, the models are in both cases off by a factor of at least 3. To illustrate this, in Figure we see the second wave in Stockholm (blue) along with two attempts to model it using the code by [12] (that was published in Science, red and yellow curves chosen to match 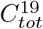 and *T*_*wave*_ respectively). The green and purple use similar codes but include pre-immunity, the details of which will be further explained in Section 3.

The remaining explanation that avoids pre-immunity is that *fluctuations* in the effective *R*-value, denoted *R*_*e*_, due to NPI’s and/or behavioral changes completely controls the way the cases evolve with time, rendering the SEIR-model useless. Yet, the NPI’s in Sweden have remained virtually unchanged and SEIR-models are not anywhere near being able to match the development in Sweden (as seen in Figure 3, yellow and red curve) so if this is true it must be caused by public awareness and voluntary behavioral changes by the population. In the coming section we take a closer look at this potential explanation.

**Figure 3:**
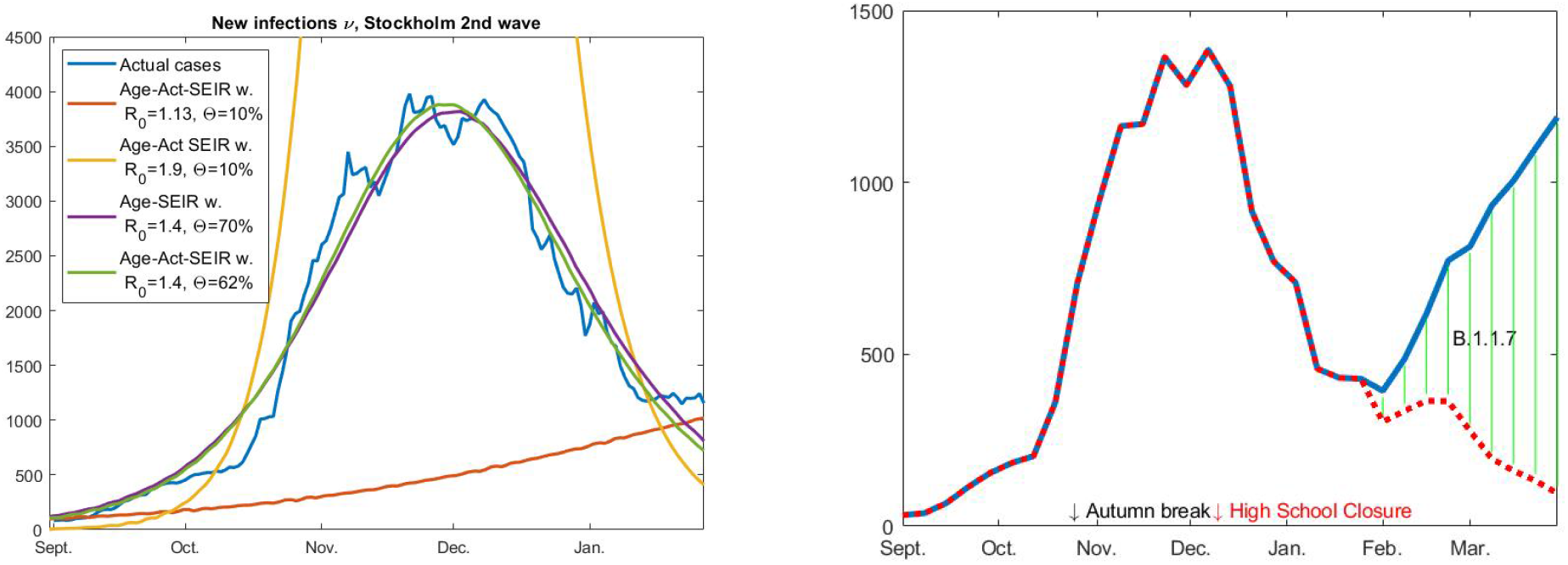
Left: Cases for Stockholm second wave (re-scaled to match an increase of 11.5% sero-prevalence) and various attempts to fit with model curves. Right: Cases (for Stockholm) continued into February and March, with and without B.1.1.7.

### 2.3 The case of Stockholm, Sweden

The second wave hit Sweden much later than many other countries, leading to wide speculation that the “Swedish strategy” had been successful. We now know it did not give us advantages for the second wave, as the Swedish Health authority had predicted. A recent study estimates that the strategy lead to between 26 to 82% unnecessary deaths and no notable improvement for other factors, such as the economy [9]. We focus our analysis on the metropolitan area of Stockholm with 2.4 million inhabitants, which has been hit by 3 distinct waves, spring and autumn 2020 and a third wave driven by the British mutation B.1.1.7 during the spring of 2021. In June 2020, 11.3% were estimated to have been infected in Stockholm, yet the decline of the spread started in early April, a month after onset of the first wave, even though the mitigation strategies were very limited and not well followed by the Swedes. From mid March, high schools and universities were on distance learning, people were expected to work from home and if possible avoid public transportation. Frequent washing of hands, keeping distance of 1.5-2 m, and staying at home if feeling sick, were the main recommendations given to the public from the health authorities and the government. A lock-down was never implemented, family members of confirmed cases were expected to work, grade 1-9 schools remained open and face masks were not recommended. It seemed odd that the soft interventions could have this effect unless the population was protected by some degree of preexisting immunity. Nevertheless, it is of course possible that these interventions along with public awareness and arrival of spring was enough to keep *R*_*e*_ below one from early April and onwards.

The same argument is however harder to accept considering the second wave (seen in Figure 3, blue curve, left as well as right). The recommendations from the government were the same as described for the first wave and public compliance limited; there were frequent reports of full shopping malls and crowded transit stations, very few people wearing masks and the weather getting colder, driving people indoors. As is clear to see in Figure 3, the spread started to dampen in late October and peaked in late November, yet the only noteworthy change in recommendations occurred on December 7 when high schools went online. Thus it is unlikely NPI’s which makes the steep rise in October to level out and then recede. It is important to realize that in the absence of the British mutation B.1.1.7 the epidemic in Stockholm would most likely have remained calm up until present at least, see the red dashed curve in Figure 3, right. In our subsequent publication [14] we look much closer on Stockholm and the argument that variations in public behavior could have caused the ups and downs, demonstrating that this explanation is unlikely.

In summary, if SEIR-models have any validity at all in describing the spread of SARS-CoV-2, we find that it is very hard to explain the second wave in Stockholm without taking pre-existing immunity into account. This indicates that a temporary herd-immunity level, given the soft restrictions, had been reached in Stockholm county in December 2020 (similar to Manaus between its first and second wave). Similar observations are also reported from Lombardy, Italy [56]. The authors write “It appears that a comparatively high cumulative incidence of infection, even if far below theoretical thresholds required for herd immunity, may provide noticeable protection during the second wave” and then argue that these observations can not be attributed to neither NPI’s nor behavioral or environmental variations alone.

## 3 SEIR-modeling with pre-immunity

Several scientists have proposed that a level of pre-immunity within the population could partially explain the unexpected behavior of the SARS-CoV-2 virus spread in society [24, 36, 40], but without providing a thorough mathematical analysis. Similar arguments have also been put forth by Doshi [19] as well as by Sette and Crotty [51], see SM Section 5.2 for a longer discussion. If a pre-existing immunity of substantial relevance would exist, it would significantly affect the model, but in this article we do not speculate in the nature of the pre-immunity, we just provide mathematical support for its existence. We refer to our subsequent publication [15] for a more profound analysis of the possible mechanisms behind this pre-immunity.

### 3.1 Incorporating pre-immunity into the SEIR model

The simplest way to incorporate pre-existing immunity in the mathematical model is to select an immunity level of Θ percent and set *s*_*initial*_ = 1 − Θ*/*100, where *s*_*initial*_ is the initial fraction of susceptible individuals in the population, (see SM Section 5.2 for details). The only assumption needed for the SEIR-model to work in theory, is that the NPI’s and public behavior remain rather constant during the elapse of the wave, and that seasonal factors remain fairly constant during the given time window. We recall that 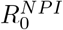 denotes the amount of people one infected individual transmits the disease to, at the onset of the epidemic with NPI’s in place, *and assuming that everyone else is susceptible, i*.*e. s*(*t*) ≈ 1. As long as NPI’s remain constant, it follows that 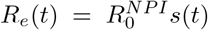. However, it is important to realize that in the presence of pre-immunity this formula needs to be reweighted, as follows

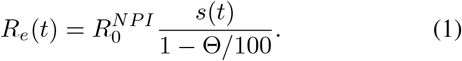

We introduce a new variable *α* for the number *R*_0_*/*(1 − Θ*/*100) which thus represents the (initial) transmission rate in a group of fully susceptible individuals, and 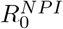 the initial transmission rate in a mixed population with a given pre-immunity level.^2^

A more refined way to introduce pre-immunity is by including variable susceptibility, since “being immune” is not a binary variable. On the contrary, neither cross-reactive adaptive nor innate immunity is likely to hinder infection when exposure to the virus is high. We describe the mathematical details in Section 7.6. In particular, we show in Section 7.7 that there is only a slight difference between modeling variable activity levels and variable susceptibility, so in practice it may be hard to distinguish one of these explanations from the other.

To exemplify this, in Figure 4 we plot the output of the basic homogenous SEIR-model (with pre-immunity Θ = 35%), the age-stratified model by Britton et al. (with pre-immunity Θ = 25%), the age-activity stratified model (again by Britton et al., now with Θ = 0% and “activity parameter” *η* = 2, see Section 6.1 or 7.7 for more details), and finally a more advanced age-susceptible stratified model where 25% of the population has no protection at all against the virus, whereas the remaining 75% with pre-existing immunity are divided into three groups and given various levels of protection. In all cases, *R*_0_ ≈ 2.

**Figure 4:**
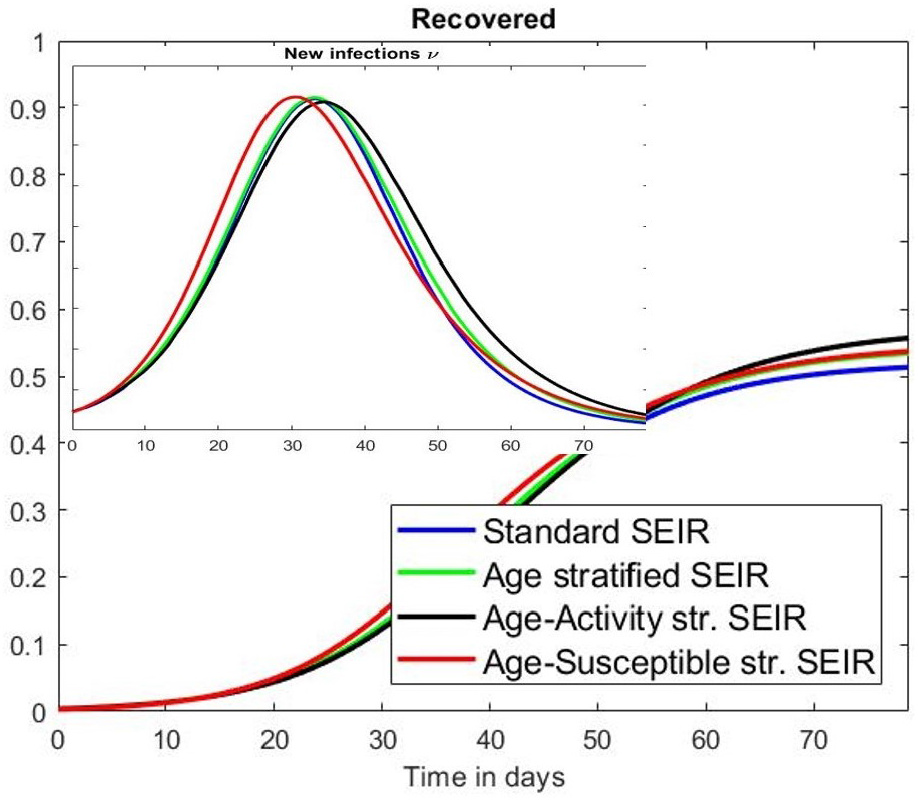
Blue: Standard SEIR (2) with pre-immunity level Θ = 35%. Green: Age-stratified SEIR with Θ = 25%. Black: Age-activity model with “activity difference factor *η* = 2” and Θ = 0. Red: Agesusceptible stratified SEIR.

As is plain to see, given any of the above SEIR-models it is possible to chose parameters in any of the other models to get almost identical output. From this we can draw a number of interesting conclusions:

1. It is impossible within this mathematical framework to draw any certain conclusions about the nature of the pandemic, since different real world phenomena have an almost identical effect on the curves. However, this conclusion only holds within a certain parameter interval; as we saw in Figure 3 it is impossible to model the development in Stockholm with the age-activity stratified model and no pre-existing immunity, without choosing absurd values for the parameter *η* regulating activity variation. This is further elaborated in SM Section 6.1.
2. Immunity is rarely either 100% or 0%, whether a person gets infected will depend on the dose of infectious virus attacking a person with different protective antibody and T-cell immunity levels. Despite this, a more refined model taking various levels of immunity into account, and a simpler model treating the biological reality as binary, give the same output. Hence, if one is careful with the *interpretation* of Θ as the amount of “immune”, it is perfectly fine from a mathematical viewpoint to replace the more advanced age-susceptibility stratified model with a more simple one. See [15] for an in depth study of this phenomenon.

### 3.2 Modeling examples with pre-immunity

We exemplify by applying the model age-stratified model by Britton et al. [12] to the situations in Manaus, India (first wave) and Stockholm (second wave), and we satisfy with modeling pre-immunity using the simpler approach with a “binary” parameter Θ for the percentage of pre-immune. In SM Section 6.1 we show that the age-activity stratified model by Britton et al. is less realistic^3^, especially for modeling poorer places like Manaus and India, but we have still chosen to include it when modeling Stockholm to demonstrate that even this model drastically fails to give a good fit with observed data, unless pre-immunity is taken into account.

We will first focus on Manaus, since this is an example of a relatively unmitigated spread that is believed to have reached saturation without seasonal interference. In Figure 5 we display the result using pre-existing immunity level Θ = 42%. The output overlaps surprisingly well with observed data, the shape and *T*_*wave*_ are reasonable and 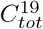 becomes 44%, which is what was measured in [13]. For this to occur we chose *α* = 3.45, which corresponds to 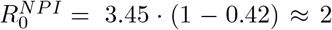. The age-distribution has been adjusted for Brazil, as explained in Section 7.1. Above we used the measured data value of 44% and not age-sex reweighted of 52% [13]. Whether we pick 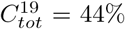 or 52% has no bearing on the major conclusions, using 52% gives 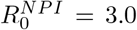 and Θ = 33%. If, as argued in [30], the population sero-prevalence was closer around 30%, then we need a pre-immunity of around 60% to get a good model fit, which is in line with our conclusions from Stockholm.

**Figure 5:**
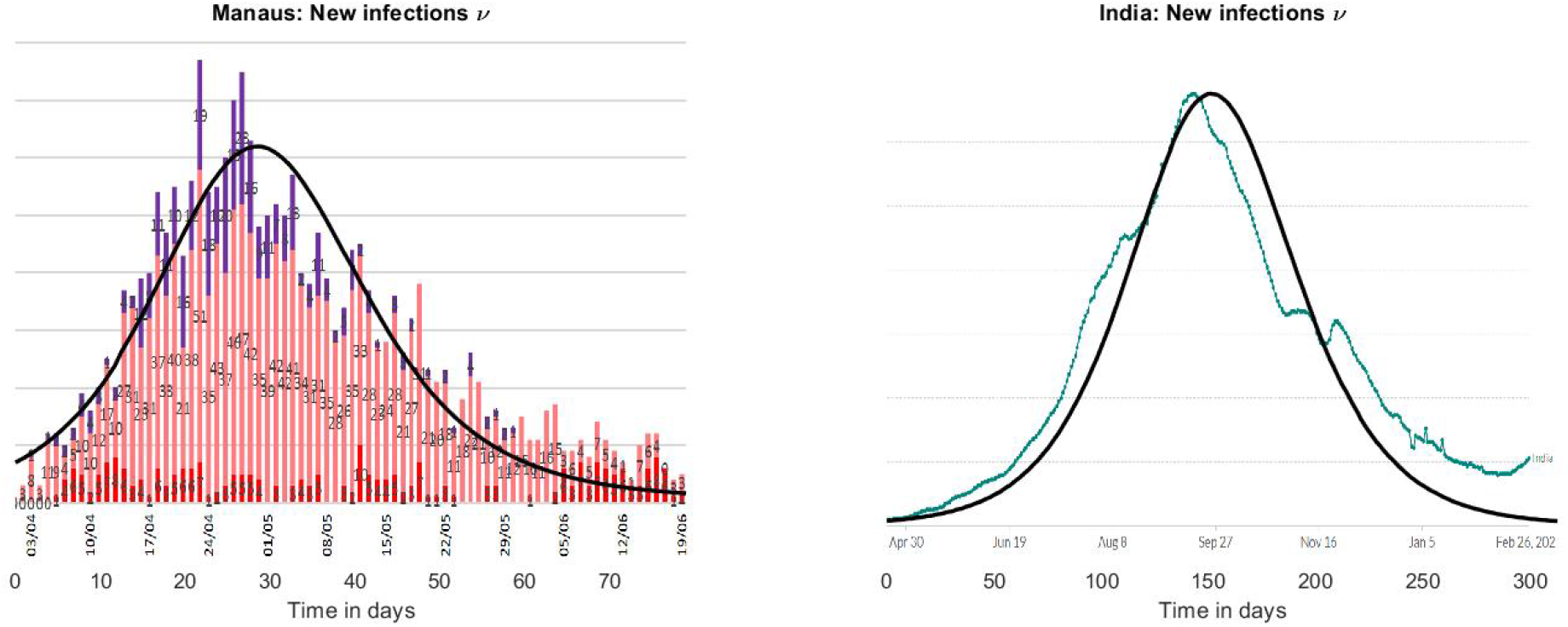
Left: Age-stratified SEIR with pre-immunity 42%, which gives 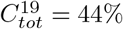. y-axis have been scaled to fit curve of deaths in Manaus. Right: Same model run for India with pre-immunity 42% (again), giving 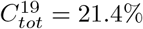. y-axis have been re-scaled.

We then ran the same model for the country of India, again setting Θ = 42% but now choosing a lower value for 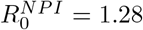 in order to get 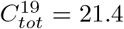, which was the national seroprevalence as estimated in the end of February by ICMR (Indian Council of Medical Research). That 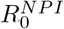 vary between countries could have a number of explanations, depending on climate, population density, NPI’s etc. India is currently seeing a substantial second wave, which is due to a more contagious mutant virus variation, and hence this does not contradict the above modeling.

Before proceeding, we would like to stress that this research is in no way meant to support mitigation strategies to control the pandemic, since the examples set by e.g. Sweden and Brazil clearly shows that this has led to substantial amount of human suffering and unnecessary loss of life [9]. Moreover the tragic second waves hitting both Manaus and India shows with all clarity that this is not a path to be considered; even if Manaus and India had “herd-immunity” for some time, the recent surges is probably related to both mutations and loss of immunity [49], and both issues speak firmly for relying on suppression/vaccination strategies [4]. On the other hand, if some degree of pre-existing immunity is present in the world’s population, we believe that this is an important fact that can be used for better prediction of the pandemic and understanding of the virus.

In fact, the new waves caused by the mutations are commonly explained by attributing to them a higher *R* − value, but the hypothesis of a pre-existing immunity offers an alternative interpretation. If there was a pre-existing immunity caused by cross-reactivity with other viruses, which according to the above modeling could have protected at least 40 percent of the population, then any mutation which is able to get past this pre-immunity will seem like it has a higher *R*, when in fact it simply has a larger pool of people it can infect. This is further investigated in [14].

### 3.3 Stockholm, second wave

We now describe in further detail our modeling of Stockholm which we already displayed in Section 2.3, Fig. 3. The Swedish second wave is a good option for mathematical modeling, as public recommendations have remained rather constant except for the closing of high-schools on December 7. Since we are modeling the second wave, it is important to have a measurement of the immunity level at the onset of the wave. Interpolating between the measurement of 11.3 and 9.6 in mid-October [23], we argue that the sero-prevalence was 10% on September first, which is the starting date in our model. Note that the recent study [17] shows a moderate decline of antibodies to SARS-CoV-2, and also the study [29] has established that prior infection results in 80% protection against reinfection, which did not seem to wane over time. Therefore we have not included loss of immunity over time in this model, due to the relatively short time window. Another quantity is the under-reporting factor in the publicly available time-series of COVID-19 cases. The sero-prevalence in Stockholm rose to 22.6% in early March [23], which is in line with an independent study [46] that concluded that 17% of Stockholm blood donors were seropositive in the midst of the second wave. Based on this, we estimate that the under reporting factor is 2.4, which corresponds to an increase in sero-prevalence of 11.5% in the actual time interval between 1/Sept/20 and 28/Jan/21. The blue curve seen in Fig. 3.3 is the seven day average of actual cases, (so 1/9 actually displays the average of 1-7/9), as reported by the Swedish Public Health Agency, multiplied by 2.4.

We tried to find parameters that would fit the shape of this time-series. In short conclusion, it is impossible to match this data with any SEIR-model not incorporating levels of pre-existing immunity above 40%. In order to get a good fit with the age-activity stratified SEIR [12], we had to choose the activity parameter *η* = 100, which is absurd. Or rather, in practice this means that the spread only takes place among the 25% “highly active”, and hence this is the same as assuming that 75% are either pre-immune or isolating, see SM Section 7.7.

We display a few of these tests in Figure 3. We first ran Britton et al’s Age-Activity stratified SEIR model with a pre-existing immunity of 10%, corresponding to pre-pandemic immunity of 0%. In order to get 11.5% as the “final size of the second wave” (corresponding to the measurement of 22.6 mentioned earlier), we needed to pick transmission rate *α* = 1.26, corresponding to 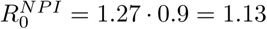, the result is seen in red. In order to get a realistic value for *T*_*wave*_ we need to pick 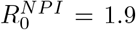, leading to the yellow curve and a final size of the second wave of 48% (compared with actual 11.5%). Looking at the corresponding curves, it is apparent that both alternatives are completely off, confirming what we have previously stated; It is impossible to get near reality, even using age-activity stratified models, without including pre-immunity.

Next we tried to find values of 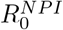 and Θ that would match 11.5% as the final size of the second wave, using the simpler Age-Stratified SEIR. The choice Θ = 70% and 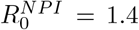 gives a perfect fit. If again we incorporate the various activity levels by Britton et al. (*η* = 2), then these values drop further to Θ = 62% and still 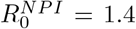. The age-activity model is more realistic in the Swedish scenario than Manaus/India, due to the social well-fare state and extensive IT-infrastructure, combined with many single households and online based jobs, which makes it easier for large portions of the population to self-isolate to various degrees. For this reason we ran both models for Stockholm, and so combined this gives evidence for a pre-existing immunity in the range 62-70%. Since this is the second wave, the values of pre-existing immunity also includes the 10% acquired during the first wave, so these values correspond to pre-pandemic immunity of 52 − 60%.

One may argue that the above figures are unreliable, since the model for example does not take into account isolation of the elderly. This is indeed the case, and for that reason we have further developed the model in [14], taking the third wave, vaccinations and isolation of elderly into account. This more advanced model puts the pre-pandemic immunity at 62%, and hence the figure is relatively stable. Note that we do not claim that this proves that the above estimates are precise, it simply shows that one need to use pre-immunity levels of above 40% to get acceptable fit between model and observation, which makes the standard hypothesis of zero pre-existing immunity very doubtful.

### 3.4 The Gompertz curve and stability of estimates

In this section we briefly discuss if the heterogeneous model can be further tuned to more accurately fit observed data, without resorting to pre-immunity, to test if such refinements could alter the above conclusions. The first waves in a number of places had the shape of a “Gompertz”-curve (characterized by a sharp rise and a slow decline). In the supplementary material, we propose a socio-economic stratified SEIR model to explain this. The idea is simple, if you have a rich part of town and a poorer part of town, then the transmission rate *α* is likely to be higher in the former and lower in the latter. Once you add up the curves, the result looks like a Gompertz-function, as shown in Figure 6 (left). We also managed to fit Manaus almost perfectly by setting the pre-existing immunity Θ = 43% (Figure 6, middle), again abiding by the restriction 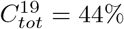 as measured in [13]. This should be compared with Θ = 42% used in Figure 5. It is remarkable that, despite variations in the model, key behavior is surprisingly consistent, and we have not been able to get a similar fit with any reasonable model and Θ = 0%, indicating a certain robustness to choice of model.

**Figure 6:**
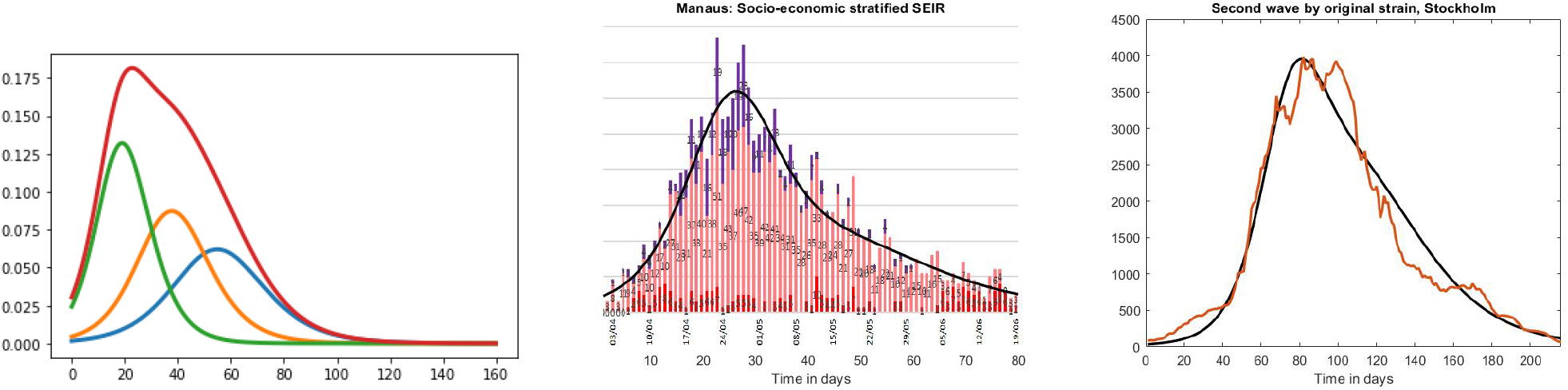
Left: 3 independent SEIR-curves for different *R*_0_-values, and their sum. Middle: Manaus modeled using Θ = 43% giving 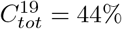. Right: Stockholm modeled using Θ = 62% giving 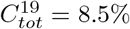.

The same comment goes for Stockholm, to the right in Figure 6 we see the second wave in Stockholm not including B.1.1.7, (compare Fig. 3). We modified the Socio-economic age-activity SEIR-model a bit further to take into account that the 70+ group is to a large extent self-isolating, and that social mixing in between age-groups certainly is reduced during the pandemic. All of these factors help to reduce the herd-immunity threshold, and hence increase the possibility of getting a good fit with observed data without using pre-immunity. The outcome is seen in black, using a pre-immunity of 52%, corresponding to a pre-pandemic immunity of 42%. In summary, we see that more advanced models can yield even better accuracy, but the pre-immunity parameter Θ remain in the same magnitude. In particular, it is still impossible to get near observed data with Θ = 0, and this gives mathematical support to the hypothesis that a certain level of pre-existing immunity was present before the arrival of SARS-CoV-2.

## 4 Conclusion

We undertook a major evaluation of different SEIR-models taking factors such as age, activity level, susceptibility variation, socio-economic level and pre-existing immunity into account. We tested these models against data acquired from Manaus, India and Stockholm, and found that a simple age-stratified SEIR-model with pre-immunity in the range 40-60% gave good fit with observed data, which is impossible to attain in the absence of pre-immunity, as the other factors can not affect the model sufficiently. We also analyzed a number of potential alternative explanations and found that they did not seem backed up by data, so we argue that if SEIR-models are at all useful for modeling COVID-19, then pre-existing immunity is a plausible explanation to the unexpected development of the pandemic that should encourage further investigation in order to reveal its nature.

## Supporting information

Supplementary Material: The mathematics of SEIR

## Data Availability

Data and code will be made available on github once the manuscript is accepted for publication.

## Acknowledgement

We thank the authors of [8] for sharing their data with us. We thank Jakob Svensson and Anders Vahlne for interesting and fruitful discussions on the subject. We thank Andrea B. Simons for help with Brazil data. We thank Cecilia Hellström, Peter Nilsson and Sophia Hober for their contribution to antibody pre-immunity data and Birger Sørensen and Andres Susrud at Immunor AS for collaborative work on identifying the Influenza A/SARS-CoV-2 cross protective immunity. We thank Afsar Rahbar, Nerea Martín Almazán, Mattia Russel Pantalone for data regarding pre-immunity serology prevalence.

## Author contribution

Concept: Cecilia Nauclér-Söderberg and Marcus Carlsson. Mathematical analysis: Marcus Carlsson. Development and testing of numerical code: Marcus Carlsson and Gad Hatem.

## Data availability

Numerical codes and relevant data-sets will be posted on GitHub upon acceptance of the manuscript.

## Footnotes

1 The measured and theoretical curves will not be equal since there is always a proportion that do not get tested. Also, to be correct, on should apply a time-shift to account for *T*_*incubation*_

2 In the first version of this paper, we defined 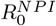 as *α/γ*, but decided to change this since it deviates too far from the accepted role of *R*_0_. See Section 5.2 for more details.

3 at least not with activity level parameter *η* = 2, and for reasonable values of *η* the difference with only age-stratification becomes negligible

