## Supplementary Material: The mathematics of SEIR for "Mathematical modeling suggests pre-existing immunity to SARS-CoV-2"

In this supplementary material we aim at explaining how to construct more advanced SEIR models in a mathematically sound way, starting from the very basic (homogenous) one, in Section 5. Section 6 explains heterogeneous models in an expository way, with a focus on how various parameters affect the dynamics of the model and why it is easy to draw false conclusions from such models. Finally, in Section 7 we give a rigorous mathematical treatment of heterogeneous SEIR-models and on how to incorporate factors such as variable infectiousness, susceptibility, social activity and socio-economic level.

### 5 Basic SEIR

We begin with introducing the SEIR-equation system in its most basic form

$$\begin{cases} s'(t) = -\alpha s(t)i(t) \\ e'(t) = \alpha s(t)i(t) - \sigma e(g,t) \\ i'(t) = \sigma e(g,t) - \gamma i(g,t) \\ r'(t) = \gamma i(g,t) \end{cases} \tag{2}$$

Here, $\alpha$ represents the rate at which infectives transmit the disease to a fully susceptible population. The appearance of $s$ on the right hand side of the equation $s'(t) = -\alpha s(t)i(t)$ represents the basic epidemiological fact that you can not infect people who are no longer susceptible (see e.g. first equation, Simple epidemics, [5]). One may improve this system in an innumerable number of ways, taking into account pre-symptomatic spread, loss of antibodies, geographical constraints etc.; we refer to [10] for a concise overview.

We first focus on describing the basic setup of the model. The parameters $\alpha$, $\gamma$ and $\sigma$ relate to $R_0$, $T_{incubation}$ and $T_{infective}$ by the formulas $\sigma = 1/T_{incubation}$, $\gamma = 1/T_{infection}$ and $\alpha = R_0/T_{infection}$. However, while estimates for $T_{incubation}$ center around 5-6 days [37, 34, 59], estimates for $T_{infectious}$ are virtually non-existent but around 5 days seems to be a good estimate [60]. Despite this, expert teams like the Imperial College group [57] and Britton et al [12] use much lower values; $T_{incubation} = 4.6$ and $T_{infectious} = 2.1$ in the former and $T_{incubation} = 4$ and $T_{infectious} = 3$ in the latter. To understand why requires some in depth understanding of their role in the above equation system, which we hereby describe.

Imagine that you were to infect the entire population at one single time $t = 0$. They will then become infective and recover at rates decided by $\sigma$ and $\gamma$. The corresponding equation system (with $s(t) \equiv 0$) is easily solved by hand; $e(t) = \mathrm{e}^{-\sigma t}$ and $i(t) = \frac{\sigma}{\sigma-\gamma}(\mathrm{e}^{-\gamma t} - \mathrm{e}^{-\sigma t})$ (where $\mathrm{e} = 2.718..$ is the natural exponent, not to be confused with $e$, which has a different font). These curves are highly counter intuitive, since the proportion of people remaining in $e$ decays exponentially, whereas making a concrete choice of $T_{incubation} = 5.7$ (from [59]), say, gives the impression that most people stay in $e$ for around 5.7 days and then move on to $i$ (see Figure 7). Let us explore to some extent the reasons for, and the implications of, this seemingly wrong nature of the equation system.

The fact that the system is set up like this reflects that it is the limit of stochastic Markov-processes, i.e. random processes with no "memory" (Ch. 5.5 [3]), and hence the system can not keep track of how long a "person" has been in a given compartment (such as $e$). The connection between $\sigma$ and $T_{incubation}$ comes from the fact that "on average", a person stays $\int_0^\infty e(t)dt = \int_0^\infty e^{-\sigma t}dt = 1/\sigma$ days in the exposed compartment. The formula $\gamma = 1/T_{infective}$ is derived similarly. In [3] this is commented as follows: "The assumption of an exponentially distributed infectious period is certainly not epidemiologically motivated, although with this assumption the mathematical analysis becomes much simpler." The authors of [10] are on the same note; "This assumption, while making the models and their analyses easier, is not biologically realistic for most infectious diseases" (Ch. 3.6). In section 4.5 and 4.6 of the same book they proceed to introduce more realistic systems of integral equations. Such systems have a long history, see e.g. [31] and [39], where it is established that these models are more sensitive, and [22] argues that the use of these more sophisticated methods can lead to different policy decisions. This raises the question;

*Why are SEIR-models still in use for modeling COVID-19?*

Before giving an answer to this question, let us further explore the counter-intuitive nature of (2). Let us suppose that the "entire population" in the above example is not the entire population but just 100 individuals who are surrounded by fully susceptible individuals. Then $I(t) = 100\frac{\sigma}{\gamma-\sigma}(\mathrm{e}^{-\gamma t} - \mathrm{e}^{-\sigma t})$ describes how many of these 100 that are infective at time $t$. The amount of spread these persons gives rise to at that moment in time then equals $\alpha I(t)$, (compare with the first equation of (2)). If we again divide by 100 we get a function which describes the rate $\beta$ at which an individual transmits

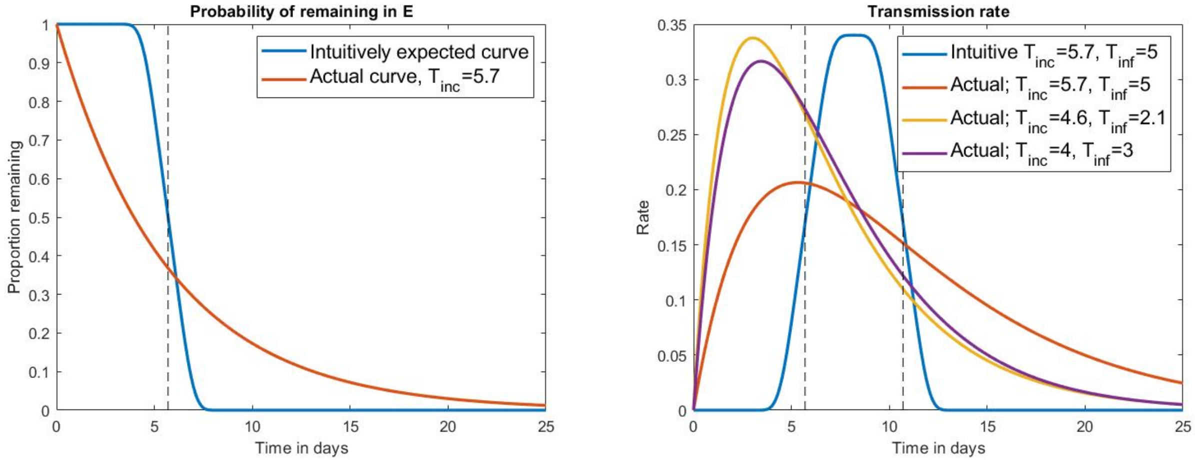


Figure 7: Left; likelihood of remaining in group $e$ (dashed line indicates $T_{infective} = 5.7$). Likelihood to transmit COVID-19 as a function of time

.

the disease, as a function of time from when that person became exposed, i.e.

$$\beta(t) = \alpha i = \alpha \frac{\sigma}{\sigma - \gamma}(\mathrm{e}^{-\gamma t} - \mathrm{e}^{-\sigma t}). \tag{3}$$

We display the transmission rate $\beta$ for various parameter choices in Figure 7. Again, the curves look nothing like one would expect based on intuition; despite an incubation time estimated to $5.7$ days, most people start transmitting the disease immediately, with the transmission rate reaching its peak even before the end of the incubation time. Hence, there is a huge gap between the mathematical models and the medical reality. Rather surprisingly, we now demonstrate that these shortcomings have an insignificant effect on modeling outcome, given a careful (and somewhat counter intuitive) parameter choice.

Integral equation versions of SEIR takes the age of infection as well as estimates of the Transmission rate into account, and are therefore much more realistic. Such equation systems, of which the standard "differential equation SEIR" is a simplification, have their roots in the original work by Kermack and McKendrick [33] from 1927, and are described e.g. in Section 4.5 of [10]. We implemented such an equation system relying on a Transmission rate almost identical to the "intuitive transmission rate" seen in Figure 7. The result is displayed in Figure 8, along with corresponding curves for the standard SEIR based on parameters $T_{incubation} = 5.7,\ T_{infectious} = 5$ as well as the Imperial College choice $T_{incubation} = 4.6,\ T_{infectious} = 2.1$. As is plain to see, there is virtually no difference between the Standard SEIR (with parameters from the Imperial College Team) and the more advanced "integral equation SEIR". Clearly, this is one answer to the question posed earlier in italic.

#### 5.1 Parameter choice

What remains unclear is why it is the Imperial College parameters that gives the best fit, rather than $T_{incubation} = 5.7,\ T_{infectious} = 5$ which the transmission rate $\beta$ was based on (the parameters used by Britton et al gives a curve somewhere in between the yellow and red). To understand why, we further explore the significance of these parameters in the mathematical model.

To derive the equation relating $\alpha$ and $R_0$, note that integration of (3) gives the total amount of reinfections that one individual gives rise to, which yields the equations

$$R_0 = \frac{\alpha}{\gamma} \Longleftrightarrow \alpha = \frac{R_0}{T_{infectious}}, \tag{4}$$

(assuming, as we do here, that there is no pre-immunity). This has a natural intuitive interpretation; the rate of transmission is the total transmission divided by amount of days a person is infectious. Similarly, for the integral equation SEIR we have $R_0 = \int \beta dt$.

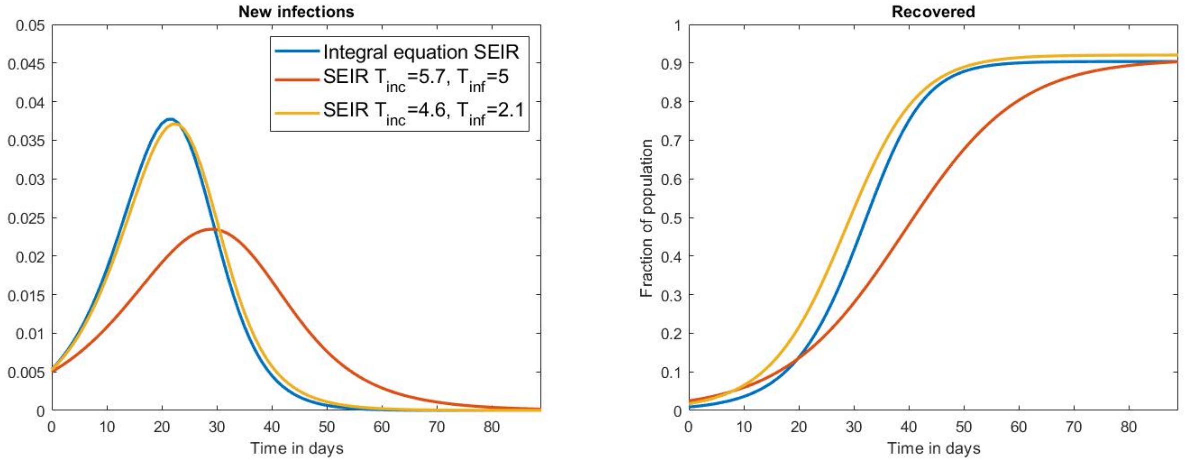


Figure 8: Curves for different models and parameter choices. $R_0 = 2.5$ is fixed.

In Figure 8 we display "the integral equation SEIR" along with the standard SEIR for two different parameter choices of $T_{infectious}$ and $T_{incubation}$, whereas we kept $R_0$ fixed at 2.5. We observe that the final size of the epidemic $C_{tot}^{19}$ depends largely on $R_0$ alone, whereas the model choice or other parameters $T_{incubation}$ and $T_{infectious}$ mainly affect the duration of the wave. We recall that the herd-immunity threshold is given by $1 - 1/R_0$ which for $R_0 = 2.5$ gives $0.6$. It is interesting to see that $C_{tot}^{19}$ becomes much larger, 0.9, which is due to the "overshoot" effect stemming from having many infected simultaneously at the peak of the wave. This gives some mathematical support for the usefulness of lockdowns, since if we stop the pandemic at $r(t) = 0.6$, and then restart it with $s(0) = 0.4$, no epidemic will unfold. Hence, given a fixed $R_0$, the final size of the epidemic range from $60\%$ to $90\%$ depending on the NPI's.

Another quantity of interest is the *serial interval* or *generation time* $T_{generation}$,[4] which is estimated to 6.7 days here [8], other references report $T_{generation} \approx 4$, see [20] and the references therein. Mathematically, the generation time can be computed by normalizing the curve in (3) (so that it represents probability density function for transmission) and then computing the expectation. This gives, as expected

$$T_{generation} = T_{incubation} + T_{infective}. \tag{5}$$

Armed with these observations, we now explain the somewhat counterintuitive parameter choices for $T_{incubation}$ and $T_{infective}$:

$T_{incubation}$; From a medical perspective, the incubation time is the time from infection to onset of symptoms, but in the SEIR model, the incubation time stops when you start to infect others. So if we believe that the real incubation time is $T_{incubation} = 5.1$ (from [37, 34]), we must remove time accounting for pre-symptomatic spread. If we estimate this to be 0.5 days, we arrive at $T_{incubation} = 4.6$ (which is the argument used in the SM of [57]).

$T_{infective}$; one would expect that the infectivity correlates with the viral load, for which there is estimated a peak around 5 days from symptom onset and very little transmission beyond 10 days (see e.g. the meta-study [60]). But this is uninteresting for the mathematical model for two reasons; First, these studies usually deal with hospitalized patients, whereas the mathematical model is concerned with the "average" person who gets COVID-19. Second, most infected will isolate themselves after symptom onset (except for family contacts), and the mathematical model is really concerned with the average time a person *transmits*, not the average time they *could potentially transmit*. Hence the use of $T_{infective}$ is misleading and should be $T_{transmissive}$. Estimates for $T_{transmissive}$ seem non-existent in the literature, which is not rare, how would one design a study measuring this in practice? Therefore, $T_{infective} = T_{transmissive}$ is chosen so that (5) is satisfied, using an estimate of $T_{generation}$ instead. We can now see where the Imperial College Team picked $T_{infective} = 2.1$ from, since $6.7 - 4.6 = 2.1$. (Luckily, they did not pick the estimate $T_{generation} = 4$ from [20], which would have put $T_{infective}$ at $-0.6$. Where Britton et al got the values $T_{incubation} = 4$ and $T_{infectious} = 3$ from is not clarified in their publication, but there is a marginal difference to the numbers 4.6 and 2.1.) Whichever choice one makes, the choice of parameters is clearly connected with great uncertainty, which is another reason for using these models with

[4]technically, the generation time (time between subsequent infections) and the serial interval (time between symptom onset) are different, as is explained e.g. here [35]. For our model we need data on the generation time but this is impossible to observe, so instead we will estimate this based on observations of the serial interval.

caution. On the other hand, as was previously mentioned, different parameter choices for $T_{incubation}$ and $T_{infectious}$ seem to yield very similar curves, which is an indication that the curves produced by SEIR may be relevant despite this flaw. We have used $T_{incubation} = 4.6$ and $T_{infectious} = 2.1$ for all plots in this paper.

Let us finally explain why the "Imperial College curve" (yellow) fits with the "Integral Equation SEIR curve" (blue) in Figure 8, as opposed to the red curve seemingly based on the same parameters as the blue! (See Figure 7, right). Arguing as before, we see that the formula for $T_{generation}$ for the integral equation SEIR becomes $T_{generation} = \int t\beta(t)dt / \int \beta dt$, which turns out to be 6.7, despite the transmission rate being chosen to intuitively give $T_{incubation} \approx 5.7$ and $T_{infective} \approx 5$. Hence the blue and yellow curve share the same $T_{generation}$-value, which is what really influences the speed with which the pandemic grows. The corresponding number for the red curve is 10.7, which is completely off.

An alternative approach to picking numbers for $T_{incubation}$ and $T_{generation}$ would be to completely ignore the medical *interpretation* of these mathematical constants, and simply try to find numbers so that the "transmission rate distribution" in Fig. 8 b) fits with corresponding curves actually estimated from medical research. For example, it is widely believed that transmission of SARS-CoV-2 happens mainly near symptom onset [47], so then the cumulative distribution of the incubation period, which is estimated in Figure 1A of Bi et al [8], should be roughly the same as the curves we get by integrating the transmission rates in Figure 8. We display the result in Fig. 9, and as is plain to see, the "yellow" parameter choice gives the best fit. Similarly, transmission rate distributions are estimated in [32] (Figure 1) and, while neither of the curves in Figure 8 give a very accurate overlap, the yellow one seem to give the best overall fit, again lending support to the choice of 4.6 and 2.1.

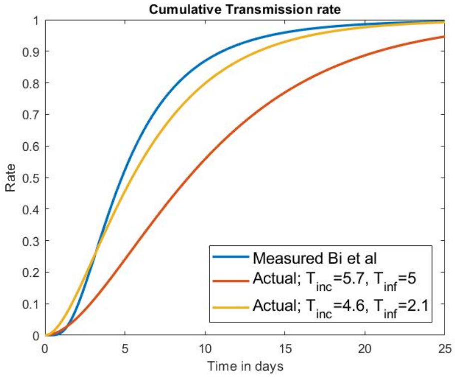


Figure 9: Fitting parameters to measured data.

But, at the end of the day, we have to admit that very little is still known about the actual shape of the transmission rate, (as argued e.g. here: [47]). Therefore, as an alternative to "best guess" parameter choice one can also do stability tests, running the same model with parameters ranging inside certain bounds, as suggested in [28], or switch to fully Bayesian probabilistic models [24, 61]. However, these models are a bit like a "black box" for non-experts, and in the light of recent developments and the rather opposite conclusions of the two papers [24, 61], both relying on "Bayesian SEIR-models", this seems to be no bullet proof way to avoid contradictory outcomes. In this article we go ahead with the "best guess", hoping to make advanced SEIR models less of a "black box" to the reader. Also, as noted above, rather different parameter choices for $T_{incubation}$ and $T_{infective}$ have very limited effect on outcome, mainly affecting $T_{wave}$, in our experience. In summary, it seems that the SEIR-model has a sort of built in robustness making output usable even if input parameters are slightly off.

#### 5.2 What is pre-immunity and how does it affect the mathematics?

The key problem addressed in this paper, i.e. the discrepancy between model and measured data, can be easily fixed by adding pre-immunity, as we now explain. First of all, if a fraction $\theta$ have pre-existing immunity to SARS-CoV-2, note that the true $R_0$-value must be much higher than the observed value. In fact, it is easy to see that

$$R_{0,observed} = (1-\theta)R_{0,true} = (1-\theta)\frac{\alpha}{\gamma}. \tag{6}$$

(We reserve the term "$R_0$" for $R_{0,observed}$, since this corresponds with the standard interpretation as well as output of $R$-estimation packages such as EpiEstim [55].) Secondly, note that if $s, e, i, r$ is a solution to (2) where $\alpha$ is given by (4), then $(1-\theta)s, (1-\theta)e, (1-\theta)i, (1-\theta)r$ is a solution to (2) with $\alpha$ given by (6). The key problem addressed in this note is that if we chose an $R_0$ such that the time $T_{wave}$ corresponds with reality, then usually the final size of the epidemic $C^{19}_{tot}$ is above 70%, i.e. completely out of bounds. The second observation means that this can be "corrected" by simply rescaling all curves to get the desired value of $C^{19}_{tot}$, upon including a corresponding level of pre-immunity.

We sum up these simple observations in a theorem.

**Theorem 5.1.** *For every solution to* (2) *in the absence of preimmunity, there is a corresponding solution in the presence of a preimmunity level $\theta$, simply obtained by rescaling the first solution by the factor* $(1-\theta)$*. In particular, the value of $T_{wave}$ remains the same whereas the new value of $C^{19}_{tot}$ becomes $(1-\theta)$ times the old value.*

For example, suppose we have found parameters such that the shape of the modeled "wave" fits well with observed data, but the magnitude is off so that $C_{tot}^{19} = 80\%$ but the measured sero-prevalence is $20\%$, it could be that the case that the model actually "works" if there is a pre-immunity of $75\%$ (since $0.2 = (1-0.75)\cdot 0.8$).

Several scientists have proposed that a level of pre-immunity within the population could partially explain the unexpected behavior of the SARS-CoV-2 virus spread in society [24, 36, 40]. Lourenco et al [40] suggested that already a 10-20$\%$ acquired immunity level to SARS-CoV-2, and a pre-existing immunity level of whatever kind, could build up to herd immunity levels to limit a virus outbreak, but without providing any analysis of the applicability of their basic "SI" mathematical model to SARS-CoV-2. In [24], a more sophisticated Bayesian approach was used to predict the spread, but since this model predicted very mild second waves, the model appears to be less trustworthy. The mathematics in [36] is rather weak, mainly pointing out that the virus spread does not seem to be exponential in many places, suggesting that Gompertz-type functions are more suitable than the curves obtained by traditional SEIR-modeling.

In this study we do not speculate further about the nature of what we call "pre-immunity", which mathematically simply means "individuals who do not seem likely to get infected even if exposed to the virus". This could be caused by pre-existing cross-reactive adaptive immunity or different degrees of innate immunity, but also people who self-isolate will be included in the pre-immune group.

Concerning pre-existing cross-reactive adaptive immunity, several studies have demonstrated that T-cells from healthy individuals respond to SARS-CoV-2 peptides in vitro, even so T-cells collected from individuals before the SARS-CoV-2 virus existed [11, 27, 41, 50]. As such, they confer a pre-existing immunity and could in theory dampen the virus-spread. It has been proposed that this memory T-cell response could be mediated by a cross-immunity to common cold corona viruses; i.e that these T-cells are trained to respond to corona viruses but due to cross immunity protect against the new SARS-CoV-2 virus. However, although T-cells are very important in the combat of this virus, these are not considered to protect against infection, but are likely more important to protect someone from becoming severely ill. Instead, antibodies rather than T-cells would be more important to prevent an airborne transmitted virus such as SARS-CoV-2 to infect another person. To our knowledge, only one study so far presented evidence for the existence of specific cross reactive antibodies to the cold coronavirus, which was found in $44\%$ of children and $5\%$ of adults [43]. These antibodies had neutralizing capacity and are hence potentially able to protect against SARS-CoV-2 infections. Since this study yields pre-immunity levels of between 40 and 60$\%$, this indicates that the virus is not yet fully understood, and calls for further research into the mechanisms behind the pre-immunity.

### 6 Heterogeneous SEIR-models, an overview

The idea of using heterogeneity to obtain more nuanced and accurate SEIR models is widespread and explained in textbooks such as [10]. For example, the Imperial College Team uses age-stratified models taking into account demographics as well as varying societal and mixing patterns [57]. In [24] the model takes into account heterogeneities such as susceptibility, transmissibility, exposure and location. A Swedish epidemiological modeling group has published a very intricate code where Sweden is divided into 21 regions [53], each region modeled by its own set of S,E,I and R-functions along with 6 additional functions representing e.g. people who need hospitalization, ICU care, number of deaths and so on. The flow of travelers between the various regions was assumed to be governed by certain laws, and moreover the various functions representing subgroups for each region were dispatched into subgroups based on age, with assumptions of inter-age mixing patterns. A similar model was proposed by Lipton et al. in a more general context [38].

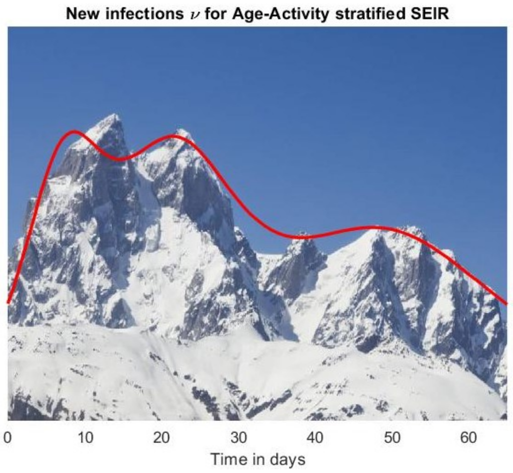


Figure 10: Mount Ushba modeled using three socio-economic compartments.

It is important to understand that for each new variable added to the equation system, there comes along mathematical constants whose values need to be guessed, since it is not possible to measure their true values. In other words, with a very complex model it is often possible to create an output that coincides with measured data, *whether or not the model actually represents reality to any degree*. To underline this point, we modeled the silhouette of mount Ushba (Svaneti region, Georgia) using three socio-economic compartments

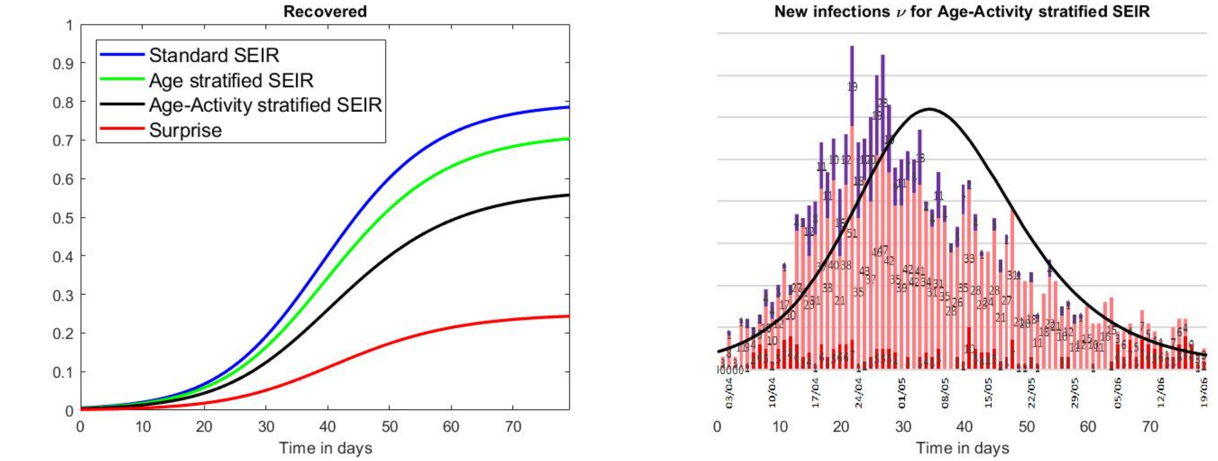


Figure 11: Left: Various levels of heterogeneity and $R_0^{NPI} = 2.0$. Right: new infections in the age-activity stratified SEIR with the curve of deaths from Manaus in the background [2]. The time axes have been synchronized.

(Section 7.8).

For a more serious example, the authors of [53] manage to fairly accurately match the Swedish deaths and ICU-occupancy during the spring of 2020. But a closer inspection of the graphs reveals that the model yields other values that are completely at odds with measured data, in that $C_{tot}^{19}$ is out of bounds, similarly to Figure 1. More precisely, the authors consider 5 scenarios labeled $a)$-$e)$ depending on the level of social distancing and separation of age groups. The computed curves for Stockholm with 2.4 million inhabitants in scenario $a) - c)$ yields $R$-curves that level out near 2.4 million, so $C_{tot}^{19}$ for Stockholm comes close to $100\%$, which is of course totally unreasonable. The best fit of both deaths and ICU occupancy happens for alternative $d)$, but in this case the epidemic ends with approximately $800000$ infected, i.e. $33\%$ of the population, see Figure 2b [53], which is off by a factor 3. Even for alternative $e)$, which includes so restrictive measures that deaths and ICU are underestimated, yields $C_{tot}^{19}$ for Stockholm of roughly $25\%$. This exemplifies that it is much easier to create SEIR models that accurately match deaths and ICU-occupancy, since these depend on unknown constants that can be chosen to fit data, whereas main output like $C_{tot}^{19}$ and $T_{wave}$ are harder to match. In other words, the problem with the behavior seen in Figure 1 and 2 seems to persist even in this very advanced model.

Another modification to the standard SEIR-model was considered by Britton et al [12], with the idea of introducing population heterogeneity as well as different social activity levels. We ran their model with a “true” value of $R_0 = 2.5$ and a reduction due to NPI’s of $\alpha$, so that $R_0^{NPI} = \alpha \cdot R_0$. In Figure 11 (left) we display the result for $\alpha = 0.8$.

As is plain to see, the age-stratification leads to a modest reduction in the final size of the epidemic ($C_{tot}^{19}$), whereas adding also variable activity levels has a more substantial impact. We also used the new infections curve $\nu$ from the Age-Activity stratified SEIR (after rescaling and time-translation) to fit with the Manaus death time series. The rationale for this is the assumption that the $\nu$-curve will be similar in shape to the deaths curve, mainly differing by magnitude and translation in time. Clearly, this model is not too far off from describing what happened in Manaus (Figure 11 right). The epidemic seems to “burn out” at around $55\%$, which roughly is the sero-prevalence level of Manaus in June according to the post-correction done in [13] (that gave $52\%$, while the measured sero-prevalence was $44\%$). So could it be that this additional complexity is the variable that other scientists overlooked in the beginning of the pandemic? We doubt this conclusion, as we explain in the coming section.

#### 6.1 Is variations in social activity enough to accurately model the epidemic?

We here provide a very brief summary of what is explained in depth in the Supplementary Material, Section 7. In the age-stratified model, each function $s$ , $e$ , $i$ and $r$ is the sum of 6 different curves; one for each of the 6 age-groups 0-5, 6-12, 13-19, 20-39, 40-69 and 70+, respectively. Each age group contains a fraction of the population $\mathbf{w} = (w_1, \ldots, w_6)$. A matrix called $A$ (see (10)) controls how many interactions there are between individuals in different age-groups and $p$ gives the probability that an interaction leads to infection. In the absence of pre-immunity these are connected with $R_0$ via the formula

$$R_0 = pT_{infectious}\rho(\mathrm{diag}_{\mathbf{w}} A) \tag{7}$$

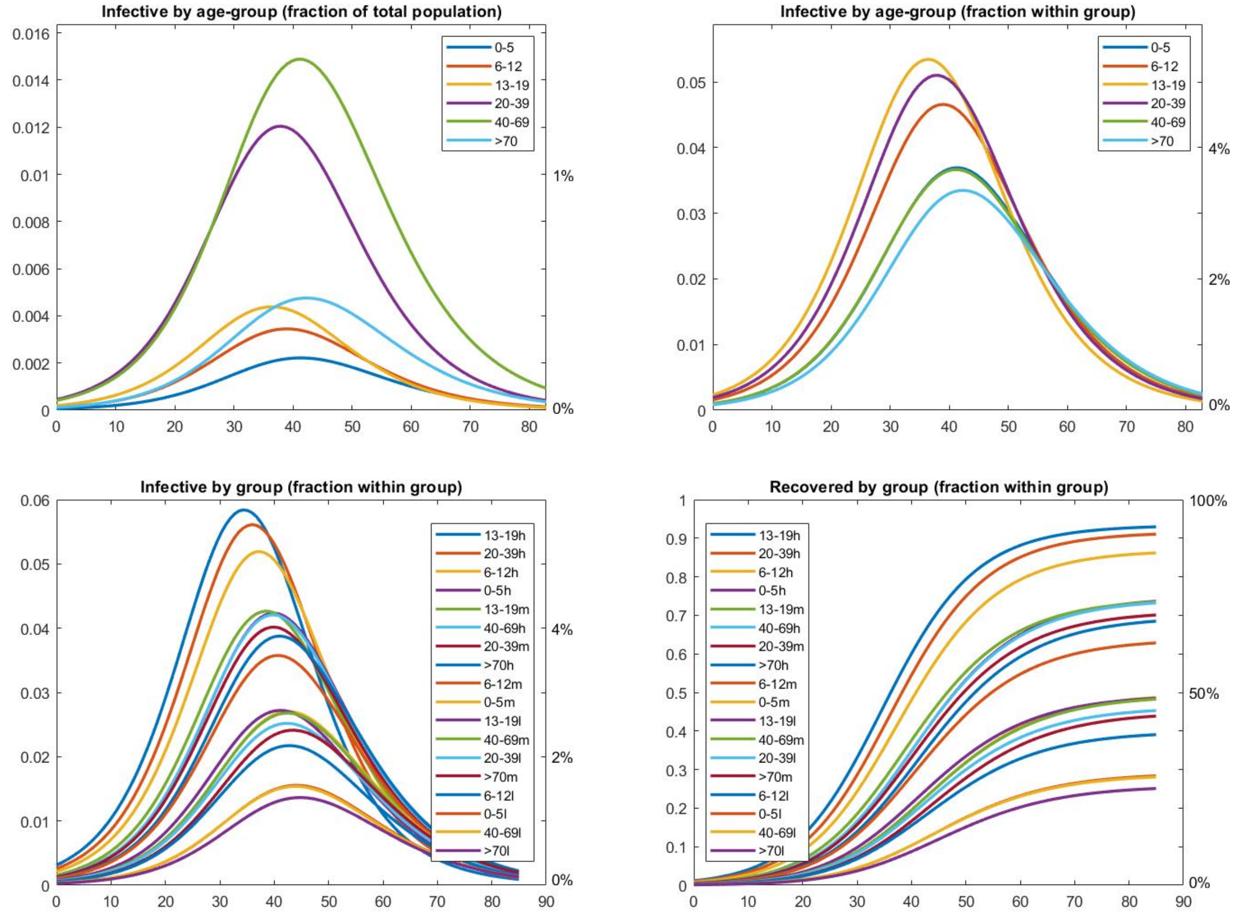


Figure 12: In all graphs, $R_0^{NPI} = 2.0$. Top panel: Age-stratified SEIR. Bottom panel: Age-activity-stratified SEIR.

where $\rho$ denotes the "spectral radius" of the matrix $\mathrm{diag}_{\mathbf{w}} A$ (compare with (4)).

To get an understanding of what this new level of complexity really does in the model, we plot in Figure 12(a) each of the 6 "$i$-functions", using the same model/parameters as for the green curve in Figure 11. Since this is not too informative, we also provide a plot that instead displays the fraction of new infections in each subgroup in (b). A clear pattern emerges; the age groups 13-19 and 20-39 appear as "the main drivers of the pandemic" (followed by age 6-12), whereas especially the group 70+ become infected to a lesser degree and somewhat later in time. These observations suggest that they primarily get infected as a consequence of the large spread in the younger groups, which is in line with observational studies such as [42]. Also, a Swedish government report conclude that the main reason for the high death tolls among the elderly in care homes in Sweden was the due to high community spread, rather than which NPI strategies and routines different elderly care homes had implemented [54].

We next incorporate activity-stratification into this model as well; each age group is now divided into subgroups; $25\%$ with low social activity, $50\%$ with medium social activity and $25\%$ with high social activity. Mathematically, low activity means that people in this group have half the amount of daily interactions with respect to the medium group, whereas the high activity group has twice as many. In Figure 12 (c-d) we run the same experiment for the age-activity stratified model, but we sorted the labels according to final size of the epidemic in each respective group. $l$ stands for "low activity", $m$ for "medium" and $h$ for "high". As is plain to see, it is, as expected, the highly active ones that are now driving the pandemic, with the same three age groups as before being in the top league, reaching final sizes of around $90\%$, whereas only $25\%$ of the low active portion of $70+$ becomes infected.

But is this a reasonable model to describe the real spread of the SARS-CoV-2 virus? Six out of the eight first places are held by the high-activity group, with the highly active 70+ group reaching an infection level of $70\%$, way ahead of the medium active 40-69 group which ends at $50\%$. It then seems as the entire pandemic is driven by the highly active group. This "feeling" can be demonstrated mathematically, by considering reducing the amount of interactions in the highly active group to normal levels (i.e. we dividing the activity level in the top $25\%$ by 2). By Theorem 7.1, this reduces the $R_0-$value from 2.0 to 1.04, which means that in practice there will be no epidemic. But is it possible that it is the social "over-activity" of only $25\%$ of Manaus inhabitants that were driving the outbreak?

Another issue with this explanation of why the epidemic would "burn out" at around $55\%$ is that division $25\%,\ 50\%,\ 25\%$ and the corresponding activity levels $1/2, 1, 2$ is clearly quite arbitrary, so we replaced this with $1/\eta, 1, \eta$ for a new parameter $\eta$. The curve labeled "surprise" in Figure 11 is the same model run with new activity levels $\eta = 10$ (as opposed to $\eta = 2$), and now the epidemic burns out at $25\%$. While this argument by itself does not disqualify the correctness of using activity stratification, it makes the model rather useless for any sort of prediction, since it is impossible to measure the "correct" value in reality.

Furthermore, Manaus is located in a relatively poor region of Brasil, with less than $25\%$ of the population having a formal employment and according to official statistics they have an average wage of 600 USD [44]. It therefore seems more likely that $75\%$ of the population would have more *similar* activity levels. This is coherent with the findings in [13] that reports similar sero-prevalence estimates across the various sub-groups studied, suggesting that the virus spread appears to have occurred relatively uniformly across the population and was not limited to specific subsets of people who may have been more exposed. Hence, we argue that the age-stratified SEIR model (green curve in Figure 11) is the most reasonable model to use for the situation in Manaus. But this model predicts final size of the epidemic to $C^{19}_{tot} = 70\%$ in June, whereas measured data from [13] was $44\%$. Admittedly, this was adjusted upwards to $52\%$ due to age and sex re-weighting, and then further adjusted to $66\%$ due to rapidly waning antibodies, but it has been argued that this is an overestimate [4] which even the authors of the original study seem to consider plausible [49]. Indeed, very reasonable arguments are put forward in [30], concluding that the true peak sero-prevalence in Manaus during the first wave probably was around $30\%$. In any case, the Manaus sero-prevalence quickly dropped to below $30\%$ and despite this, the epidemic remained calm until a more contagious variant started a second wave. This behavior is not expected if the age-activity stratified SEIR was a good model for COVID-19 spread in Manaus.

Finally, looking at how well the model can fit observed data for Stockholm in Fig. 3 (yellow and red curves), we draw the conclusion that variations in social activity is far from enough for accurate modeling COVID-19.

### 7 The mathematics of heterogeneous SEIR-models

We first introduce the Age-Activity-Susceptibility-Stratified SEIR model by Britton et al [12], and then explain how to nuance this to take other factors into account. We model the spread of the disease with a multidimensional compartment model with functions $S, E, I$ and $R$, which will depend on 3 "group"-variables plus time:

$g_1 \in \{1, \ldots, 6\}$ - age group. We let the amount in group $g_1$ be $N_{g_1}$, so that $N = \sum N_{g_1}$. The fraction of individuals in each group is then determined by the function $w_{age}(g_1) = N_{g_1}/N$.

$g_2 \in \{1, \ldots, n_{act}\}$ - activity level. The weight function $w_{act}$ determines what portion (of each age group) are in respective "activity-group". The activity level is decided by a function $f_{act}$ where $f_{act}(g_2) = 1$ corresponds to the subgroup $g_2$ which has normal infectivity level, $f_{act}(g_{act}) = 2$ means they have twice that of normal etc.

$g_3 \in \{1, \ldots, n_{sus}\}$ - susceptibility level. This is similarly determined by a function $f_{sus}$ where $f_{sus}(g_3) = 1$ corresponds to normal infectivity level and $f_{sus}(g_3) = 0$ indicates total immunity. As before this is accompanied by a weight function $w_{sus}$ which determines the fraction of the population in each respective group.

Thus, there will be a total of $N_G = \{1, \ldots, 6\} \times \{1, \ldots, n_{act}\} \times \{1, \ldots, n_{sus}\}$ subgroups, each determined by a tuple $g = (g_1, g_2, g_3)$. For each such group there will be one function $S(g, t)$ representing the actual amount of susceptible individuals in group $g$ at time $t$, whereas $s(g, t)$ is defined as their fraction *in the total population*: $s(g, t) = S(g, t)/N$. Similarly we have functions $E,\ e,\ I,\ i,\ R,\ r$ each depending on $g$ and $t$. The total fraction $w$ in a given group $g = (g_1, g_2, g_3)$ is then

$$w(g) = w_{age}(g_1) w_{act}(g_2) w_{sus}(g_3) \tag{8}$$

and in a more refined setup we could drop the tensor structure and just let $w$ be a non-negative function on the set of possible tuples with $\sum_{g \in G} w(g) = 1$. This would allow for e.g. children to be less susceptible to the virus than adults, etc. Similarly, $f_{sus}$ could also depend on e.g. age group $g_1$, but we do not investigate such refinements here.

#### 7.1 The contact matrix

To model how the disease spreads within and between the different groups, we need some sort of measure of how often these groups interact. Wallinga et al [58] divided in 2006 the Dutch population into 6 groups aged 0-5, 6-12, 13-19, 20-39, 40-59 and 60+ and measured the amount of average weekly conversations held between group $j$ and group $g$. To be precise, Wallinga et al presents a "contact matrix" $M$, which is a $6 \times 6$ matrix whose $j, k$:th entry equals the amount of conversations between age group $j$ and $k$ on a weekly basis, *divided by* the amount of people in group $k$. Hence $M_{j,k}$ is the average number of persons in group $j$ that a person of group $k$ has contacts with in a week (based on data from the Netherlands in 2006).

Several issues arise when modeling COVID-19 using this matrix; the age distribution in e.g. Brasil is clearly different than that of the Netherlands and furthermore, even the Netherlands has a very different age distribution in 2020 than in 2006. For example, in [58] it is stated that the age group 60+ is about 17%, whereas the age group 70+ in 2020 make up $14\%$, due to an aging population. For modeling of COVID-19 it is clearly better to have 70 as a delimiter for the last age group, since 70+ is the group that many countries have tried to protect extra from the infection. We thus need to decide on how to update $M$ and $A$ given a new age distribution $w_{age}^{new}$.

Recall that the elements of $M$ represent "the average number of persons in group $j$ that a person of group $k$ has contacts with". If the age groups change a bit or delimiters are moved, it is reasonable to assume that these numbers are mildly affected, so let us agree that they should be constant. Now $M_{j,k}w_{age}^{new}(k)N$ is then the amount of conversations between groups $j$ and $k$, whereas by reciprocity $M_{k,j}w_{age}^{new}(j)N$ represents the same number, but these two numbers will no longer be the same. To remedy this issue, we suggest to define $M_{j,k}^{new}$ by taking the average and then divide by the amount of people in group $k$, i.e. $w_{age}^{new}(k)N$. This leads to the formula

$$M_{j,k}^{new} = \frac{M_{k,j}w_{age}^{new}(j) + M_{j,k}w_{age}^{new}(k)}{2w_{age}^{new}(k)}. \tag{9}$$

We now drop the sup-index "new", assuming this has been cleared, and proceed to introduce the matrix

$$A_{j,k} = \frac{M_{j,k}}{7w_{age}(j)}$$

which thus is a symmetric matrix (since $M_{j,k}w_{age}(k) = M_{k,j}w_{age}(j)$), where the factor 7 is convenient, since it allows us to talk of daily contacts instead of weekly. For applying these matrices to other countries, an obvious objection is that people in e.g. Brazil are more social and live in households with more members, why all numbers should be increased with some factor. On the other hand one can argue that they should be decreased by some factor due to NPI's. Thus, let $\mu$ be a correction factor so that the actual amount of encounters is $\mu A$. Another observation to take into account is that during NPI's, contact patterns change. For example, if we want to include in our model that the elderly has been isolated more than the average citizen with some factor $\xi$, we can do so by multiplying each element of the last row and column by $\xi$. To illustrate using the data from [58], we have

$$A(\xi) = \begin{pmatrix} 24.2 & 4.5 & 2.5 & 4.9 & 2.3 & 1.6\xi \\ 4.5 & 39.2 & 4.6 & 5.0 & 2.9 & 1.6\xi \\ 2.5 & 4.6 & 32.0 & 7.2 & 5.4 & 2.1\xi \\ 4.9 & 5.0 & 7.2 & 10.8 & 7.1 & 3.6\xi \\ 2.3 & 2.9 & 5.4 & 7.1 & 8.8 & 4.7\xi \\ 1.6\xi & 1.6\xi & 2.1\xi & 3.6\xi & 4.7\xi & 7.7\xi \end{pmatrix}. \tag{10}$$

Summing up, the daily contacts will be described by the matrix $\mu A(\xi)$ and, unless explicitly stated, we assume that $\mu = \xi = 1$ and write $A$.

We use this matrix as a measure of how much the different groups are in contact with each other, although we are aware that this matrix is suboptimal in many ways for modeling COVID-19. First, the age distribution as well as contact patterns certainly vary between countries. Secondly, the grouping of all individuals above 60 is very unfortunate, since it is primarily the group over 70 that have been isolated during the pandemic (due to their higher mortality), while 60-69 to a large extent are still working. We therefore postulate that the matrix would be rather similar *if the 5th group included 40-69 and the last group 70 plus*, in accordance with the previous discussion on aging population.

In Figures 5 and 6 we have updated the $A$ matrix according to formula (9) to the actual population distribution of Brazil, with the addition that the last group represents 70+. In the remaining graphs we use the Dutch population as of 2020 (which is similar to that of Sweden), again with the addition that the last group represents 70+. However, we observed no noteworthy difference due to these updates.

#### 7.2 The heterogeneous SEIR differential equations

As already mentioned, we set $s(g,t) = \frac{S(g,t)}{N}$, $i(g,t) = \frac{I(g,t)}{N}$ and so on. Let $p$ be the probability of transmission occurring when an infectious person meets a fully susceptible person and set $\beta = p\mu$. As explained above, $M_{g_1,h_1}$ is the amount of weekly conversation between an individual in group $h_1$ with persons in group $g_1$. We now refine this to consider amount of weekly conversations from a person in age-activity-susceptibility group $h = (h_1, h_2, h_3)$ with people in another group $g = (g_1, g_2, g_3)$.

If we for the moment assume that the activity-level is the same in all groups, then the amount of conversations between a person in $h$ with people in group $g$ becomes $w_{act}(g_2)w_{sus}(g_3)\mu M_{g_1,h_1}$, since $w_{act}(g_2)w_{sus}(g_3)$ is the fraction of the subgroup $(g_1, g_2, g_3)$ in the group with age index $g_1$. We now let $f_{act}$ be a vector with boost/damp-factors depending on which activity level $g_2$ and $h_2$ signify, and then we postulate (following Britton et. al.) that the amount of conversations is

$$f_{act}(h_2)f_{act}(g_2)w_{act}(g_2)w_{sus}(g_3)\mu M_{g_1,h_1}.$$

Now, we also add that people belong to variable susceptibility groups, which we model by saying that the probability of transmission during a conversation with people in susceptibility-group $g_3$ equals $pf_{sus}(g_3)$, as opposed to just $p$. Hence $f_{sus}(g_3) = 1$ means fully susceptible, whereas lower values indicate some level of protection (due to e.g. vaccination, innate immunity, cross-reactive pre-immunity etc) in the corresponding subgroup $g_3$.

At the onset of the epidemic ($t = 0$), all the persons belonging to group $g$ are in the compartment $s$, that is, they have not yet had COVID-19 so $s(g, 0) = w(g)$. In this case the amount of transmission from an infective person in group $h = (h_1, h_2, h_3)$ to group $g = (g_1, g_2, g_3)$ on a given day $t$ is

$$pf_{sus}(g_3)f_{act}(h_2)f_{act}(g_2)w_{act}(g_2)w_{sus}(g_3)\mu M_{g_1,h_1} = \beta f_{sus}(g_3)f_{act}(h_2)f_{act}(g_2)w_{act}(g_2)w_{sus}(g_3)M_{g_1,h_1}.$$

As time goes on the relative amount of susceptible in this group at time $t$ is given by $s(g,t)/w(g)$, and hence the actual amount of (weekly) transmission instead becomes

$$\beta f_{sus}(g_3)f_{act}(h_2)f_{act}(g_2)w_{act}(g_2)w_{sus}(g_3)M_{g_1,h_1}\frac{s(g,t)}{w(g)} = \beta f_{sus}(g_3)f_{act}(h_2)f_{act}(g_2)\frac{M_{g_1,h_1}}{w_{age}(g_1)}s(g,t).$$

If we divide this by 7 we get the daily amount of transmission, so upon recalling the definition of $A$ the daily transmission from one infective individual in group $h$ to group $g$ becomes $\beta f_{sus}(g_3)f_{act}(h_2)f_{act}(g_2)A_{g_1,h_1}s(g,t)$. Finally, there are $Ni(h,t)$ infective individuals in subgroup $h$ at time $t$ so the total daily transmission from $h$ to $g$ becomes

$$\beta N f_{act}(g_2)f_{sus}(g_3)f_{act}(h_2)A_{g_1,h_1}s(g,t)i(h,t).$$

Hence, introducing

$$\nu(g,t) = \sum_{h\in G}\beta f_{act}(g_2)f_{sus}(g_3)f_{act}(h_2)A_{g_1,h_1}s(g,t)i(h,t) \tag{11}$$

(i.e. the new cases in group $g$ coming from all groups $h$, divided by $N$) we see that the equation for $s$ becomes

$$s'(g,t) = \frac{S'(g,t)}{N} = -\nu(g,t), \tag{12}$$

and the remaining three "eir"-equations are then

$$\begin{cases} e'(g,t) = \nu(g,t) - \sigma e(g,t) \\ i'(g,t) = \sigma e(g,t) - \gamma i(g,t) \\ r'(g,t) = \gamma i(g,t) \end{cases}$$

The interpretation of $\sigma$ and $\gamma$ as $\sigma = 1/T_{incubation}$ and $\gamma = 1/T_{infectious}$ is the same as before. To relate $\beta$ to the previously used $\alpha$ in the homogenous model, recall that $\alpha$ is the *transmission rate*, whereas $p$ is the probability that a contact leads to infection. Hence, in the homogenous case, $\mu M$ (which then equals $\mu A$) is a number describing the rate of interaction between individuals (i.e. the amount of different individuals a person interacts with on an average day), then we would have

$$\alpha = \beta M = \beta A \tag{13}$$

where again $\beta = \mu p$. The matrix $\beta A$ can now be considered as a generalization of the number $\alpha$ to the heterogeneous setting. We usually let $\beta$ be determined by fixing a value for $R_0$, as opposed to guessing particular values. However, to explain the interrelation between $\beta$ and $R_0$ is more intricate, and done in Section 7.5.

The initial condition for this system is naturally $s(g,0) = w(g)$, $e(g,0) = \frac{n}{N}w(g)$ and $i(g,0) = r(g,0) = 0$ where $n << N$, which represents $n$ exposed individuals introduced at time $t = 0$. As long as $n$ is very small in comparison to $N$, it does not matter if we start with $n$ exposed or $n$ infective individuals, nor does it matter the distribution over the various subgroups, which we explain further in Section 7.4.

Incorporating a pre-immunity level $\theta$ in the model is a piece of cake from a mathematical perspective, we just update the initial condition for $s$ to

$$s(g,0) = (1-\theta)w(g). \tag{14}$$

This serves well to describe the effect of pre-immunity mathematically. In reality, being immune is not a binary issue, so a level of susceptibility ranging from full ($f_{sus} = 1$) to completely immune ($f_{sus} = 0$), may be more realistic, which we show how to implement in Section 7.6 (see also Figure 4, which shows that this additional complexity adds little in practice, or the subsequent article [15] which explores this in depth).

#### 7.3 Super-spreaders

With the current emphasis on super-spreaders, where 80% of the spread being attributed to around 20% of infectives [1, 21], it is of course natural to add this parameter in a similar fashion as variable susceptibility. Super-spreaders and superspreader events clearly have an impact in reality for the COVID-19 pandemic. It is therefore interesting to note that, as long as increased infectivity is not correlated with susceptibility, adding variable infectivity does not in anyway alter the total curves for $s$, $e$ etc. (obtained by summing over all sub-groups). Hence, at least in this mathematical framework, the effect of super-spreaders averages out and is therefore already built in to the $\beta$-value, describing the *average* probability of transmission of an infective with a susceptible, which we explain in depth in section 7.6. Note however that this conclusion is not necessarily true if one allocates super-spreaders randomly, we do not follow up on this here but recommend the very interesting article [25] for more on this topic.

#### 7.4 Understanding the differential equation system

We now explore the SEIR heterogeneous model a bit further. Recall that $N_G = 6n_{act}n_{sus}$ denotes the cardinality of $G$. Upon ordering the multi-index set of all possible $g$'s lexicographically, we may identify each of the four functions $s$ , $e$ , $i$ and $r$ with a (column) vector-valued function of time, to be precise an $\mathbb{R}^{N_G}-$valued function, which we shall denote by $\mathbf{s}$ , $\mathbf{e}$ , $\mathbf{i}$ and $\mathbf{r}$. For example, if both $n_{act}$ and $n_{sus}$ equal 1, then $\mathbf{s}(t)$ would be an $\mathbb{R}^6$ vector where $\mathbf{s}_1(t) = s(1,t)$ is the fraction of kids aged 0-5 who are susceptible (at time $t$), $\mathbf{s}_2(t) = s(2,t)$ the fraction in age group 6-12 etc. If we introduce two activity levels, $n_{act} = 2$, then $\mathbf{s_1}(t)$ represent the fraction of kids aged 0-5 who have low activity (corresponding to $s(1,1,t)$), $\mathbf{s}_2(t)$ represent the fraction of kids aged 0-5 who have high activity (corresponding to $s(1,2,t)$), and $\mathbf{s}_3(t)$ will be the low activity fraction in age group 6-12 (corresponding to $s(2,1,t)$) and so on.

This technical twist turns our equation system into a standard ODE, which allows us to express e.g. (11) using matrix-calculus and it also makes computer-implementation more efficient (albeit less transparent). For example it allows us to express the operation (11) as

$$\boldsymbol{\nu}(t) = \beta \mathrm{diag}_{\mathbf{s}(t)}\mathbf{A}\mathbf{i}(t) \tag{15}$$

where $\mathbf{A}$ is an $N_G \times N_G$-matrix whose entries only depend on $f_{sus},\ f_{act}$ and the contact matrix $A$. (In fact, if we omit the subgroups for activity level and susceptibility, $\mathbf{A} = A$.)

To get a hands on feeling for what this means, let us discretize the equation (12) in the simplest possible way, by letting $t$ be an integer variable describing the spread on a specific *day*, and approximating $\mathbf{s}'$ by $\frac{\mathbf{s}(t+1)-\mathbf{s}(t)}{1}$. The equation (12) then becomes

$$\mathbf{s}(t+1) = \mathbf{s}(t) - \beta \mathrm{diag}_{\mathbf{s}(t)}\mathbf{A}\mathbf{i}(t), \tag{16}$$

which is easily implemented with only basic programming skills. The "eir"-equations following (12) are equally to implement and hence the whole system can be easily computed. This simple time-stepping method (called the forward Euler method) yields graphs which are almost identical with those obtained by using more advanced ODE-packages, like ODE45 in MatLab or ODEINT in Python. In fact, judging by the supplementary material of [57], this is the method used by the Imperial College Team.

To understand how the disease progresses in the early stages, we replace $\mathbf{s}(t)$ by $(1-\theta)\mathbf{w}$ (where $\mathbf{w}$ is the vectorization of the fractions in each group according to $w$ as defined in (8)), which is a valid approximation in the early stages of the pandemic. The equations for the disease compartments $\mathbf{e}$ and $\mathbf{i}$ (analogous to (16)) can then be arranged in vector form as follows

$$\begin{bmatrix} \mathbf{e}(t+1) \\ \mathbf{i}(t+1) \end{bmatrix} = \begin{bmatrix} (1-\sigma)I & \beta \mathrm{diag}_{(1-\theta)\mathbf{w}}\mathbf{A} \\ \sigma I & (1-\gamma)I \end{bmatrix} \begin{bmatrix} \mathbf{e}(t) \\ \mathbf{i}(t) \end{bmatrix} \tag{17}$$

where $I$ denotes the identity matrix. The above is what is called power iteration, and it is well known that the largest eigenvalue/eigenvector of the above matrix determines the progression, (assuming that the largest eigenvalue is unique). More precisely, the normalized updates $[\mathbf{e}^T, \mathbf{i}^T]^T / \|[\mathbf{e}^T, \mathbf{i}^T]^T\|$ (where $T$ denotes matrix transpose) converge to the largest eigenvector, and hence the distribution in the various sub-compartments will soon resemble that of the largest eigenvector. As long as the second largest eigenvalue is not very close to the first, this process is rapid which is the reason why we claim that the initial conditions become irrelevant for the overall behavior of the modeled curves. We do not intend to quantify the above claim but simply state that in our experience this is usually the case. The amount of iterations needed for the epidemic to grow from say, 100 infected to 1000, is more than enough to make the first eigenvector completely dominant. The largest eigenvalue also clearly indicates the rate at which cases grow daily, but the formal definition of the famous $R_0$-value turns out to be connected to a closely related matrix, as we describe next.

#### 7.5 The next generation matrix and $R_0$

We now derive the relationship between $\beta$, $\mathbf{A}$ and $R_0$, following Sec 5.2 [10], and in order to save space we adapt their notation (see in particular Example 1). For simplicity, we do not make a distinction here between $R_0^{NPI}$ and $R_0$. As in the previous section we are interested in the equations for the disease compartments $[\mathbf{e}^T, \mathbf{i}^T]^T$ when $t \approx 0$. We identify one matrix $F$ responsible for the reinfections and one matrix $V$ responsible for disease progression. Upon linearizing the system (15) in the disease free case, we get (recall (14)) that

$$F = \begin{pmatrix} 0 & \beta \mathrm{diag}_{\mathbf{s}(0)} \mathbf{A} \\ 0 & 0 \end{pmatrix} = \begin{pmatrix} 0 & \beta \mathrm{diag}_{(1-\theta)\mathbf{w}} \mathbf{A} \\ 0 & 0 \end{pmatrix}$$

whereas $V = \begin{pmatrix} \sigma I & 0 \\ -\sigma I & \gamma I \end{pmatrix}$. For small $t$ our differential equation system is approximately

$$\frac{d}{dt} \begin{bmatrix} \mathbf{e} \\ \mathbf{i} \end{bmatrix} = (F - V) \begin{bmatrix} \mathbf{e} \\ \mathbf{i} \end{bmatrix}, \tag{18}$$

which is basically just restating equation (17) but in the form of a differential equation rather than recursive equation. In other words, discretizing (18) with step-length of one day will give us (17). The next generation matrix, denoted $K_L$, is now defined as

$$K_L = FV^{-1} = \begin{pmatrix} \frac{1}{\gamma} \mathrm{diag}_{(1-\theta)\mathbf{w}} \beta \mathbf{A} & \frac{1}{\gamma} \mathrm{diag}_{(1-\theta)\mathbf{w}} \beta \mathbf{A} \\ 0 & 0 \end{pmatrix}.$$

Loosely speaking, the next generation matrix is the matrix one would use if we were to model the initial epidemic growth by a power iteration using one generation as the step length, as opposed to one day in (17). The $R_0$ value is then defined as the modulus of the largest eigenvalue of this matrix, also known as the spectral radius $\rho(K_L)$. Using the well-known formula $\rho(K_L) = \lim_{n \to \infty} (\|K_L^n\|)^{1/n}$, it is easy to see that

$$\rho(K_L) = \rho(\frac{1}{\gamma} \mathrm{diag}_{(1-\theta)\mathbf{w}} \beta \mathbf{A}) = (1-\theta) \beta T_{infectious} \rho(\mathrm{diag}_{\mathbf{w}} \mathbf{A})$$

and hence it follows that

$$R_0 = (1-\theta) \beta T_{infectious} \rho(\mathrm{diag}_{\mathbf{w}} \mathbf{A}). \tag{19}$$

We now provide insights to the number $\rho(\mathrm{diag}_{\mathbf{w}} \mathbf{A})$, with a particular focus on how the introduction of new complexity levels (like adding variable activity/susceptibility) affects this value.

**Theorem 7.1.** *Consider the age-activity-susceptibility heterogeneous SEIR-model from Section 7.2. Let $A$ denote the matrix* (10) *and let $\mathbf{A}$ be as in Section 7.4. Then*

$$\rho(\mathrm{diag}_{\mathbf{w}} \mathbf{A}) = \left( \sum_{g_2=1}^{n_{sus}} w_{sus}(g_2) f_{sus}(g_2) \right) \left( \sum_{g_1=1}^{n_{act}} w_{act}(g_1) f_{act}^2(g_1) \right) \rho(\mathrm{diag}_{w_{age}} A)$$

*Proof.* For simplicity we first assume that $n_{sus} = 1$ so that the model only has two "dimensions", one for age and one for activity level, and lets denote the corresponding "$\mathbf{A}-$matrix" by $\mathbf{A}_2$. Note that $w_{age}$ and $w_{act}$ are functions, but we will without change of notation also consider them as vectors. Denote by $\mathbf{w}_2$ the vectorization of the tensor-product of

$w_{age}$ with $w_{act}$ (as in Section 7.4), and note that it takes the form of a Kronecker product $\mathbf{w}_2 = w_{age} \otimes w_{act}$. In a similar manner it is easy to see that $\mathbf{A}_2 = A \otimes (f_{act}^T f_{act})$, ($T$ denoting transpose), and moreover that

$$\text{diag}_{\mathbf{w}_2}\mathbf{A}_2 = (\text{diag}_{w_{age}}A) \otimes (\text{diag}_{w_{act}} f_{act}^T f_{act}) = (\text{diag}_{w_{age}}A) \otimes ((w_{act} \odot f_{act})^T f_{act}),$$

where $\odot$ denotes Hadamard multiplication (i.e. multiplying the elements coordinate-wise). Now, the eigenvalues of a Kronecker product between two matrices is the product of the corresponding eigenvalues, and the eigenvectors consist of Kronecker products of eigenvectors for each of the two matrices. Clearly, $((w_{act} \odot f_{act})^T f_{act})$ is a rank-one matrix whose only eigenvector is $w_{act} \odot f_{act}$, with corresponding eigenvalue $\sum_{g_1=1}^{n_{act}} w_{act}(g_1) f_{act}^2(g_1)$. Since the spectral radius is the maximum modulus of the eigenvalues, we get

$$\rho(\text{diag}_{\mathbf{w}_2}\mathbf{A}_2) = \left(\sum_{g_1=1}^{n_{act}} w_{act}(g_1) f_{act}^2(g_1)\right) \rho(\text{diag}_{w_{age}}A), \tag{20}$$

as desired.

Now, to lift this result to the full generality of three dimensions, let us denote the "three-dimensional" $\mathbf{A}$-matrix by $\mathbf{A}_3$ and the corresponding weight vector by $\mathbf{w}_3$. It is then easy to see that $\mathbf{w_3} = \mathbf{w}_2 \otimes w_{sus}$ but the matrix corresponding to $f_{act}^T f_{act}$ is now $f_{sus}^T \mathbf{1}$, where $\mathbf{1}$ is a row-vector of length $n_{sus}$. By analogous computations as above it follows that

$$\text{diag}_{\mathbf{w}_3}\mathbf{A}_3 = (\text{diag}_{\mathbf{w}_2}\mathbf{A}_2) \otimes ((w_{sus} \odot f_{sus})^T \mathbf{1}).$$

Now, $(w_{sus} \odot f_{sus})^T \mathbf{1}$ is a rank one matrix with eigenvector $w_{sus} \odot f_{sus}$ and eigenvalue $\sum_{g_2=1}^{n_{sus}} w_{sus}(g_2) f_{sus}(g_2)$. The desired identity now easily follows by repeating the argument leading up to (20).

□

We can now prove the claims made in Section 6.1 about how the $R_0$-value changes if the "highly active" group reduces their social activity by half. We had $w_{act} = (0.25, 0.5, 0.25)$. Now set $f_{act,2} = (1/2, 1, 2)$ and $f_{act,1} = (1/2, 1, 1)$. The former then gives a contribution of $0.25 \cdot (1/2)^2 + 0.5 \cdot 1^2 + 0.25 \cdot 2^2 = 1.5625$ to the $R_0$ value whereas the latter gives $0.25 \cdot (1/2)^2 + 0.5 \cdot 1^2 + 0.25 \cdot 1^2 = 0.8125$. This is a reduction by a factor $0.52$ so if the original $R_0$-value was 2 the reduced on will be $2 \cdot 0.52 = 1.04$.

##### 7.6 The effect of variable susceptibility and infectivity

As mentioned in 7.3, introducing variable infectivity does not affect the overall spread. We first explain why by using a simple example, (rather than a classical theorem/proof argument in full generality.)

Suppose that there are no age groups, or other groups either so we are back to the scalar case. Then $A$ equals $M$; the average amount of daily contacts an average person has, and $\beta$ is the proportion of those that leads to infection. With $\alpha = \beta M$ we thus retrieve the original scalar system (2) (as previously noted in (13)).

Now lets split the population in one highly infective group ($g = 1$) and a less infective group ($g = 2$). Say the first group comprises 20% and that they are 10 times more efficient in spreading the virus. Following the logic leading up to (11) we then have $f_{inf}(1) = 10$ for the super-spreader group whereas $f_{inf}(2) = 1$ for the "normal group". We then get $\mathbf{w} = (0.2, 0.8)$ whereas $\mathbf{A} = \begin{pmatrix} 10 & 1 \\ 10 & 1 \end{pmatrix} M = (1\ 1)^T(10\ 1)M$. Now lets return to the equation for disease progression (16) and suppose that $t$ is small so that $\mathbf{s}(t) \approx \mathbf{w} = (0.2, 0.8)$ is a valid approximation. The amount of daily spread in the first group is then

$$\beta M \mathbf{w}_1 \langle (10, 1), \mathbf{i} \rangle = \beta M \mathbf{w}_1 \langle \mathbf{f}_{inf}, \mathbf{i} \rangle$$

and in the second $\beta M \mathbf{w}_2 \langle \mathbf{f}_{inf}, \mathbf{i} \rangle$. In other words, the amount of spread in the two groups equals a constant times the vector $\mathbf{w}$. This means that any initial distribution of $\mathbf{i}(0)$ will soon become irrelevant and all functions $\mathbf{s}(t)$, $\mathbf{e}(t)$, $\mathbf{i}(t)$ and $\mathbf{r}(t)$ will be well approximated by $s(t)\mathbf{w}$, $e(t)\mathbf{w} \ldots$, where $s, e \ldots$ are scalar functions. These scalar functions will then satisfy a standard SEIR-equation system (2), where for example (12) turns into

$$s' = -2.8\beta M s(t) i(t) = -2.8\alpha s(t) i(t),$$

since $\langle \mathbf{f}_{inf}, \mathbf{w} \rangle = 2.8$. The only difference is that a constant of 2.8 has now appeared, an unwanted artefact that makes $\beta$ loose its interpretation as transmission probability within a contact event (cf. (13)).

So why then does variable susceptibility lead to different outcome? Again, let there be only two groups; the first group (of 20%) only gets infected with 10% probability compared with the "normal group". The matrix $\mathbf{A}$ in this case becomes

$\mathbf{A} = \begin{pmatrix} 0.1 & 0.1 \\ 1 & 1 \end{pmatrix} M = (0.1\,1)^T(1\,1)M$ and now we can observe a different amount of spread to the first group relative to the second. Given that $\beta$ is held constant, this leads to an overall slowdown of disease progression, as is to be expected.

To sum up the conclusions of this section, variable infectivity is modeled by matrices $\mathbf{A}$ of the form $\mathbf{1}^T\mathbf{f}_{inf}$, whereas variable susceptibility is modeled by $\mathbf{f}_{sus}^T\mathbf{1}$. The latter affects the overall spread, the former does not, except possibly by changing the overall rate of transmission by a constant factor.

#### 7.7 Variable activity levels

Let us now examine how variable activity, as introduced in Section 7.2, affects the model. For simplicity we assume that $n_{sus} = 1$, i.e. that the entire population is equally susceptible. Following the same logic as in the previous section, we see that a model with only activity stratification gets an $\mathbf{A}$-matrix of the type $\mathbf{f}_{act}^T\mathbf{f}_{act}$, and if there are different age-groups these matrices show up as blocks in a larger Kronecker product matrix $\mathbf{A}$.

*Thus variable activity is a bit like a cross-over between variable susceptibility and variable infectivity.* The column-vector $\mathbf{f}_{sus}^T$ is important for dividing the spread unequally, whereas the row-vector $\mathbf{f}_{sus}$ at most has the effect of adding a multiplicative constant; *Mathematically there is almost no distinction between variable susceptibility and variable activity*. In other words, if we manage to build a mathematical model based on variable susceptibility that accurately fits with an observed data collection, then it will be possible to reach the same result using variable activity-levels as well.

In order to tell which interpretation is the correct one, or rather, which is the dominant one, we need to look at what values of the involved constants are reasonable, and this requires us to first make sure we add variable activity levels in a way that unwanted constants like 2.8 in the previous section does not alter the meaning of e.g. $\beta$, $R_0$ etc. Returning to the basic construction of our equations in Section 7.2, we see that the amount of conversations between groups $g = (g_1, g_2)$ and $h = (h_1, h_2)$ before introducing the boost/damp factor $f_{act}$ equals $w_{act}(g_2)(M_{g_1,h_1}/7)w(h)N$, (assuming $n_{sus} = 1$ for simplicity) which must be interpreted as either increasing or decreasing contacts between $h$ and $g$. The total amount of conversations with these factors present is thus

$$f_{act}(h_2)f_{act}(g_2)w_{act}(g_2)(M_{g_1,h_1}/7)w(h)N. \tag{21}$$

Recalling that $w(h) = w_{age}(h_1)w_{act}(h_2)$ and that $(M_{g_1,h_1}/7)w_{age}(h_1)N$ is the total amount of conversations between age-groups $g_1$ and $h_1$, we can now sum up all the conversations between the sub-groups (21) and get the identity

$$(M_{g_1,h_1}/7)w_{age}(h_1)N = \sum_{g_2,h_2} f_{act}(h_2)f_{act}(g_2)w_{act}(g_2)(M_{g_1,h_1}/7)w_{age}(h_1)w_{act}(h_2)N$$

(since the total amount of conversations should be invariant). Upon cancelation of terms we arrive at

$$1 = \sum_{g_2,h_2} f_{act}(h_2)f_{act}(g2)w_{act}(h_2)w_{act}(g_2),$$

which in turn implies the identity

$$\sum_{g_2} f_{act}(g_2)w_{act}(g_2) = 1. \tag{22}$$

As long as this restriction is met, we can now introduce variable levels of activity while at the same time keeping disease related constants such as $\beta$ and $R_0$ fixed. For the examples in the main text we have chosen $w_{act} = (0.25, 0.5.0.25)$ and, given an "activity-difference" parameter $\eta$ (usually equal to 2), we set

$$f_{act} = \frac{1}{0.25/\eta + 0.5 + 0.25\eta}(1/\eta, 1, \eta),$$

in accordance with (22).

If a certain parameter choice gives output that fit well with reality, we can then ask the question: are the parameters reasonable? This is pursued in Section 3.3, where we argue that variable susceptibility is a more realistic explanation for the damped disease progression than variable activity. This is based on the observation that in order to get curves that fit well with data from Stockholm and Manaus, the $\eta$-parameter needs to be set unrealistically high. To be precise, we only claim that variable activity alone is not a likely explanation for the unexpectedly slow spread of COVID-19. Of course, in reality we may have a bit of both.

#### 7.8 The Gompertz curve and socioeconomic factors

COVID-19 curves, in the absence of NPI's, tend to have a "Gompertz-like" shape, i.e. a sharper rise followed by a slow decline, as noted e.g. in [36, 45]. This Gompertz-shape is especially clear in the fatality-curves for both Manaus, New York and Stockholm (during the first wave). It is unsatisfactory that none of the curves produced so far have this structure, indicating that something is missing for the model to accurately describe the reality. We argue here that socio-economic factors could be a missing piece to this puzzle. The idea is simple, imagine that you have a poor part of town, a middle class part and a rich part of town. It could be argued that, especially during a pandemic, there is very little interaction between the three groups, and that the ability to isolate oneself becomes substantially better with higher socio-economic status, both due to a lower amount of household members as well as the ability to work from home.

Mathematically, this is easily modeled by running three independent SEIR models (with whatever additional substructure) using three different $R_0$-values, and then adding the result up (with weights according to the proportion in each group). The result clearly depicts as a "Gompertz"-type curve, as illustrated in Figure 6 (left). The data from both Stockholm and Manaus supports the conjecture that this could, at least partially, explain the Gompertz-shape; in Stockholm it was the low-income neighborhoods of Tensta that got hit first in the first wave, whereas rich neighborhoods downtown had the majority of infections during the second wave. Exceptions however included wealthy areas in Stockholm with many families who had visited the Italian Alps for skiing vacation during the school holidays the last week in February, and who were not recommended to quarantine upon their arrival home. This high import of cases led to local spread of SARS-CoV-2 to similar levels as in the exemplified low income area Tensta, where few cases led to rapid spread most likely due to more social interactions. It is interesting to note that the second wave of neither Stockholm nor New York has a Gompertz shape, possibly explained by a higher level of aquired sero-prevalence in the lower socio economic classes. Indeed, a report by Public Health England [48] supports this hypothesis by the observation that the risk of contracting COVID-19 decreases with higher socio-economic level.

Similarly, in Manaus there are two major ethnical groups; whites and pardo (mixed ethnicity), plus various minor groups (black, asian, indigenous). The white population makes up for $27\%$ of the population according to Wikipedia. In spite of lack of hard facts to back this up, we now assume that these $27\%$ are better off from a socio-economically point of view. In this case, the theory we put forward here is coherent with measurements reported in the Manaus sero-prevalence study [13]; In figure S3 the monthly sero-prevalence is divided according to ethnicity, and whilst the sero-prevalence between whites and pardo show similar levels before and after the wave, in the May-measurement (mid-pandemic), whites had about half the sero-prevalence of pardo.

To build a more realistic model for varying socio-economic factors, we introduce $n_{s-e}$ socio-economic groups and an $n_{s-e} \times n_{s-e}$ contact matrix $C$, whose diagonal values represent the contact rates for each group, and whose off diagonal values describe the degree of connections between the respective groups. An age and socio-economic stratified SEIR model is then easily built by subdividing each age-group in $n_{s-e}$ subgroups and letting $\mathbf{A}$ (the matrix controlling inter-group contacts, see Section 7.4) be the tensor product of the age-contact matrix $A$ and $C$, in an analogous fashion with the previous two sections.

In Figure 6, we model Manaus using two groups (where the first group, constituting $60\%$, has a $50\%$ higher $R_0$-value than the remaining $40\%$, and with a weak coupling between the two groups). Using a (joint) $R_0$-value of $4.2$ and a pre-immunity level of $42\%$, we now obtain an almost perfect fit and a final size of the epidemic $C_{tot}^{19}$ of $44\%$, in line with the measurement in [13].

These observations support our theory that pre-immunity levels likely play a major role on the viral spread of SARS-CoV-2 and how the pandemic has evolved/unfolded. However, we again want to stress that advanced models should be used with caution, the fact that output coincides with measured data does not imply that the interpretation is correct.
